# Healthy longevity from incidence-based models: More kinds of health than stars in the sky

**DOI:** 10.1101/2021.04.16.21255628

**Authors:** Hal Caswell, Silke F. van Daalen

## Abstract

**Background:** Healthy longevity (HL) is an important measure of the prospects for quality of life in ageing societies. Incidence-based (cf. prevalence-based) models describe transitions among age classes and health stages. Despite the probabilistic nature of those transitions, analyses of healthy longevity have focused persistently on means (“health expectancy”), neglecting variances and higher moments.

**Objectives:** Our goal is a comprehensive methodology to analyse HL in terms of any combination of health stages and age classes, or of transitions among health stages, or of values (e.g., quality of life) associated with health stages or transitions.

**Methods:** We construct multistate Markov chains for individuals classified by age and health stage and use Markov chains with rewards to compute all moments of HL.

**Results:** We present a new and straightforward algorithm to create the multistate reward matrices for occupancy, transitions, or values associated with occupancy or transitions. As an example, we analyse a published model for colorectal cancer. The possible definitions of HL in this simple model outnumber the stars in the visible universe. Our method can analyse any of them; we show four examples: longevity without abnormal cells, cancer-free longevity, and longevity with cancer before or after a critical age.

**Contribution:** Our methods make it possible to analyse any incidence-based model, with any number of health stages, any pattern of transitions, and any kind of values assigned to stages. It is easily computable, requires no simulations, provides all the moments of healthy longevity, and solves the inhomogeneity problem.

## 1 Introduction

### 1.1 Longevity and healthy longevity

Healthy longevity is an inclusive term for a class of health metrics that combine information on the duration of life and on health status (Caswell and Zarulli, 2018). There is a large and increasing number of indices of the duration and quality of life, including health expectancy (HE), health-adjusted life expectancy (HALE), disability-free life expectancy (DFLE), quality-adjusted life years (QALY), disability-adjusted life years (DALY), empowered life years (ELY), and others (Robine et al., 1999; Murray et al., 2000; Jagger and Robine, 2011; Siegel, 2012; Lutz, 2017).

Two issues of increasing concern invite even greater attention to healthy longevity: population ageing and health inequality. The ageing of populations poses economic concerns that are intimately tied to the health of older individuals (Christensen et al., 2009; Institute of Medicine and National Research Council, 2013; World Health Organization, 2015; Sanderson and Scherbov, 2019), including attempts to tie retirement ages to remaining healthy longevity (e.g., van der Mark-Reeuwijk et al., 2019, in the Netherlands). Socioeconomic inequality (e.g., in income, wealth, education) has dramatic effects on life expectancy and it is important to know the impact of those same inequalities on health and healthy longevity (Jagger and Robine, 2011; Marmot et al., 2020).

Healthy longevity must be defined in relation to some type of health status. It need not refer to health conditions that are pleasant or desirable. Indeed, for many purposes, it is important to know how much life will be spent in conditions that are neither pleasant nor desirable, but which are essential components of health and which have implications for, e.g., planning for treatments or facilities. We will see examples of this in a model for colorectal cancer in Section 6.

Healthy longevity combines information on health status and age. Time spent in a specified set of health stages, or transitions among specified health stages, can have very different implications depending on the ages at which it happens (e.g., working ages, ages eligible for health insurance, fertile ages for women). The combination of health status and age can produce a truly astronomical number of possible definitions of healthy longevity (as we will see in Section 5.1, the possibilities in even a simple model dwarf the number of stars in the sky). The results in this paper give complete flexibility in choosing a definition of healthy longevity appropriate for the needs of the investigator.

The goal of this paper is a comprehensive theory and methodology for calculating healthy longevity from incidence-based health models. The method is easily and efficiently computable, extremely flexible, and does not require simulation. It provides not only the mean, or expectation of healthy life, but also the variance, skewness, and higher moments of healthy life as measures of variation among individuals.

We are concerned with four questions, the details of which will become clearer in what follows.

1. How much life will be spent in a specified health condition, over a specified set of ages? Health can be defined, as desired, in terms of disability, disease, stages of disease, need for medical care, or in other ways.
2. What is the value of the longevity in a specific health condition[s], where value may be measured by, e.g., medical costs, quality of life, or labor force participation?
3. How often, over a lifetime, will an individual make specified transitions among specified sets of health conditions? The transitions may, for example, include diagnosis with a disease or entry into or exit from institutional care.
4. What is the value of the transitions among specified sets of health conditions? Value may be measured, e.g., in terms of economic costs and benefits, types of health care required, or emotional stress.

### 1.2 Longevity as a random variable

Longevity, the duration of life (healthy or otherwise), is a random variable, because individuals are subject, throughout their lives, to probabilistic transitions among health states and from living health states to death. Probabilistic transitions imply a distribution of healthy longevity among a set of individuals experiencing a given set of demographic rates. The literature on healthy longevity focuses almost exclusively on the mean of this distribution (health expectancy), to the exclusion of the variance and higher moments. This focus is needlessly limiting. Variance has already become a recognised component of studies of longevity per se, and patterns and trends of variance (or related statistics, such as life disparity, Gini coefficients, etc.) in the age at death are well documented (e.g. Edwards and Tuljapurkar, 2005; Vaupel et al., 2011; van Raalte et al., ^2018, 2020^). The same attention needs to be paid to inter-individual variation in healthy longevity.

The need to do so is not merely academic; the variances and higher moments of longevity are important because we do not live or make decisions in a world of averages. In the context of economic planning, demographic forecasts, or policy decisions, accounting for risk is critical and requires a stochastic analysis.1

### 1.3 Prevalence and incidence

Analyses of healthy longevity can be based on the *prevalence* of health conditions or on the *incidence* of transitions among health conditions. The most common prevalence-based analysis is the Sullivan method (e.g., Sullivan, 1971; Jagger et al., 2006; Siegel, 2012), which uses the age-specific prevalence of a health condition to weight the probability of survival through an age interval, and computes the mean time spent in that health condition over the lifetime. It is widely used; in a review by Jagger and Robine (2011) of chronological series of health expectancies published since 2000, 28 out of 30 studies used the Sullivan method. Caswell and Zarulli (2018) recently presented a replacement for the Sullivan method that extends the analysis of prevalence data to include variances and higher moments.

Prevalence-based analyses have well known limitations, because they do not keep track of individuals (e.g., Rogers et al., 1989; Murray et al., 2000; Siegel, 2012). As a result, health status at one age conveys no information about health status at the next age. The prevalences of an easily reversible health condition and a permanent, irreversible condition are treated the same. Because individuals are not followed, survival is treated as independent of health status.

These limitations are relaxed in incidence-based models, which are based on rates of individual transition among health states. In incidence-based models, health status at any age can depend on health status at previous ages, and survival can depend on both age and health status. The rates of transition generally depend on age and may be functions of other covariates. Incidence-based models require longitudinal information on individuals, which is why they are so much more demanding of data, and consequently more rare, than prevalence-based models.

Incidence-based models can be formulated as multistate absorbing Markov chains^2^ with a special form, which we develop in Section 3. This formulation turns healthy longevity into a problem of occupancy time: how much of a life is spent occupying some particular set of health conditions. Because incidence-based models imply the probabilities of all the possible trajectories an individual can follow, they contain the information needed to calculate how much time an individual will spend occupying any set of ages and health conditions of interest.

The calculation of the statistics of occupancy times for healthy longevity poses a mathematical challenge, because, as we will see, occupancy times must be defined over sets of states, not single states. We solve this problem using recent developments in the theory of Markov chains with rewards (MCWRs). The resulting method is (a) powerful enough to give all the moments of healthy longevity, (b) computationally simple, requiring only straight-forward matrix multiplication, (c) flexible enough to account for occupancy defined in any way and also, for transitions between health conditions, and (d) easily generalised to attach values (e.g., quality of life) to occupancy and transitions.

#### Organisation of the paper

In Sections 2 and 3 we present the basic Markov chain framework for multistate incidence-based models that include both health stages and ages. Section 4 introduces the fundamental concept of a “reward” with which to measure (all the moments of) healthy longevity. This section includes the key methodology: the construction of the health occupancy matrix from which the rewards are calculated. In Section 6 we apply the method to a model for colorectal cancer by Wu et al. (2006). In Section 7 we summarise the results in a step-by-step protocol for the calculation of the statistics of healthy longevity. We conclude in Section 8 with a discussion the model and directions for extensions.

#### Matrices

Our analysis is formulated in terms of matrices. Matrix methods have a long history in demography, starting with early work of Keyfitz (1968); Goodman (1969); Rogers (1968) and continuing to the present. Matrix formulations have important advantages over other methods; they greatly simplify calculations that are inherently multidimensional, eliminating almost all need for the complicated summations appearing in life table methods. They yield expressions that are easily computable in matrix-oriented programming languages (e.g., Matlab and, to a lesser degree, R), and they provide access to powerful mathematical tools. The potential of matrices to unify demography was powerfully stated by Nathan Keyfitz some 40 years ago (see Appendix A).

#### Notation

Matrices are denoted by upper case bold characters (e.g., **U**) and vectors by lower case bold characters (e.g., ***π***). Vectors are column vectors by default; the row vector **x**^T^ is the transpose of the column vector **x**. The vector **1** is a vector of ones, and the matrix **I** is the identity matrix. When necessary, subscripts are used to denote the size of a vector or matrix; e.g., **I**_*ω*_ is an identity matrix of size *ω × ω*. Matrices and vectors with a tilde (e.g., **Ũ**) are block-structured, jointly classifying individuals by age and stage. The symbol *◦* denotes the Hadamard, or element-by-element product (implemented by .* in Matlab and by * in R). The symbol *⊗* denotes the Kronecker product^3^. The vec operator stacks the columns of a *m × n* matrix into a *mn ×* 1 column vector. The notation ‖**x**‖ denotes the 1-norm of **x**. Unless specified otherwise, logarithms are natural, and moments are moments around the origin, not central moments.

## 2 Markov chain models for healthy longevity

Markov chains describe the transitions of individual entities (people, for example) among states. States may be transient or absorbing. A transient state is destined to be left eventually. An absorbing state, once entered cannot be left. In our application, the transient states will be living states and the absorbing states will correspond to death. There may be multiple absorbing states if death is classified by age at death, cause of death, or other ways of leaving the population permanently.

We write the transition matrix of the absorbing Markov chain, in column-to-row orientation, as

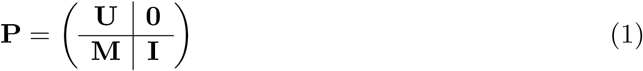

The matrix **U** contains probabilities of transition among the transient states; the (*i, j*) entry of **U** is the probability of transition from transient state *j* to transient state *i*. The matrix **M** contains probabilities of transition from transient to absorbing states; the (*i, j*) entry of **M** is the probability of transition from transient state *j* to absorbing state *i*. The identity matrix **I** and the zero matrix **0** indicate that individuals cannot return from absorbing to transient states. The entries of **P** are non-negative and the columns sum to 1.

The transition matrix **P** captures the dynamics of the probability that an individual will be in a specified state at *t* + 1, conditional on its state at *t*. Let ***π***(*t*) be a probability vector (non-negative, and entries sum to 1) where *π*_*i*_(*t*) is the probability that the individual is in state *i* at time *t*. Then

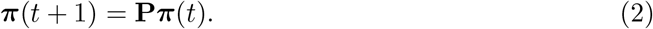

In an absorbing Markov chain, eventual absorption (i.e., eventual death) is certain. Before absorption an individual will occupy various transient states for various lengths of time. These occupancy times prior to absorption define the longevity of the individual. Expressions for the statistics (means, variances, moments) of occupancy times and times to absorption are well known in Markov chain theory (e.g., Kemeny and Snell, 1976; Iosifescu, 1980).

### 2.1 From continuous to discrete Markov chains

Individual trajectories can also be modelled in continuous time; i.e., with age a continuous variable. Some analyses of event history data, where the times of transitions are known exactly, do so naturally (e.g., Lievre et al., 2003; Cai et al., 2010; Jackson et al., 2011; van den Hout et al., 2019; Bijwaard, 2014; Brouard, 2019). See Andersen and Keiding (2002, 2015) for a survey of the statistical procedures.

Whereas the entries of **P** are probabilities of transition from one state to another over a discrete unit of time and age, the continuous time chain is defined in terms of continuous rates of transition and ageing. The dynamics are specified by an *intensity matrix*

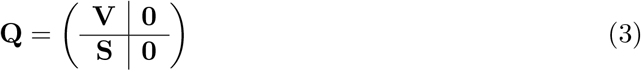

The matrix **V** contains instantaneous rates of transitions among the transient states; the matrix **S** contains rates of transition from the transient to the absorbing (dead) states. The intensity matrix **Q** has non-positive entries on its main diagonal, and non-negative entries elsewhere, and its column sums are zero.

A model with discrete health states and continuous age variation is an inhomogeneous continuous-time Markov chain, for which no solution is available. Thus it is essential to discretise the model for analysis. If **Q** can be treated as constant over a time interval (*t, t* + Δ*t*), then the discrete transition probability matrix over that interval is

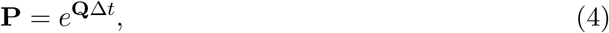

where *e*^**Q**^ is the matrix exponential function (not to be confused with the matrix containing the exponentials of the individual entries). **P** is called the discrete skeleton of the continuous model by Iosifescu (1980). This formulation requires only the that **Q** is constant between *t* and *t* + Δ*t*; choosing a sufficiently small time step will make this as true as desired.^4^

The computation of the matrix exponential is a notoriously tricky piece of numerical analysis (Moler and Van Loan, 2003). However, it is now easy to compute in Matlab, using the command expm, and the command with the same name in R.

## 3 Multistate Markov chains: combining age and health status transitions

Incidence-based models classify individuals jointly by age and health status, which we refer to in general as “health stage”. The rates of mortality may depend in an arbitrary fashion on age and health stage, and individuals may move among stages (i.e., change their health status) in arbitrary ways. The result is a multistate, absorbing Markov chain. There are many examples, including models for cardiovascular disease (Peeters et al., 2002; Ieva et al., 2017), cognitive impairment (Hale et al., 2020; Tyas et al., 2007), dementia (Coley et al., 2015; Zhou et al., 2016), disability (Mehta and Myrskylä, 2017), diabetes (Kuo et al., 1999), cirrhosis (Jepsen et al., 2015), healthy working life (Parker et al., 2020), lung cancer (Uyl-de Groot et al., 2006), colorectal cancer (Wu et al., 2006), and cervical cancer (Zhang et al., 2014).

Although presented in a dizzying variety of notations and formats, these models actually share a common underlying framework. By explicitly specifying that framework — health stages, health stage transitions, age transitions, and mortality — the construction becomes clear and analyses of explicit age- and stage-dependent results are made easier (Caswell et al., 2018).

### 3.1 Constructing the multistate model

The components of the model are a set of transition matrices among health stages with transition probabilities specific to each age class, and an age-advancement matrix that moves surviving individuals to the next older age class. The multistate model is constructed from these components using the vec-permutation matrix formalism, described in detail by Caswell et al. (2018).

It helps to begin by carefully defining some aspects of the model.

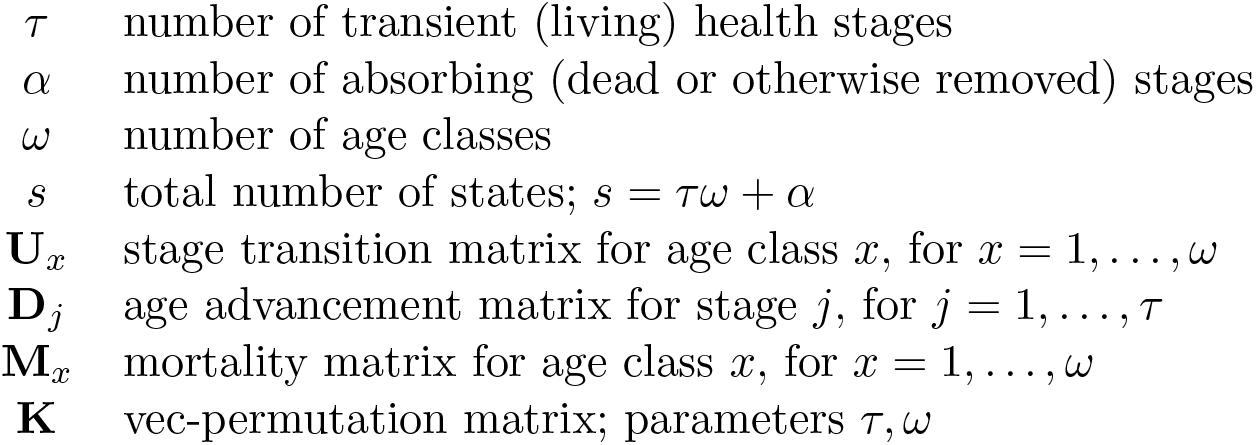

|  |  |
| --- | --- |
| $\tau$ | number of transient (living) health stages |
| $\alpha$ | number of absorbing (dead or otherwise removed) stages |
| $\omega$ | number of age classes |
| $s$ | total number of states; $s = \tau\omega + \alpha$ |
| $\mathbf{U}_x$ | stage transition matrix for age class $x$, for $x = 1, \dots, \omega$ |
| $\mathbf{D}_j$ | age advancement matrix for stage $j$, for $j = 1, \dots, \tau$ |
| $\mathbf{M}_x$ | mortality matrix for age class $x$, for $x = 1, \dots, \omega$ |
| $\mathbf{K}$ | vec-permutation matrix; parameters $\tau, \omega$ |

The matrix **U**_*x*_ describes transitions among the health stages for individuals in age class *i*. The structure of **U**_*x*_ is specific to the health condition under consideration; an example will be given in Section 3.2.

The age advancement matrix **D**_*j*_ moves surviving individuals to the next oldest age class. For example, if *ω* = 3, then

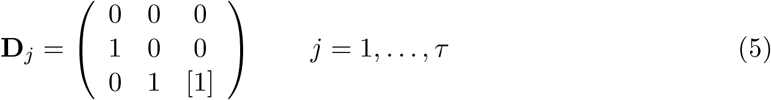

The (*ω, ω*) entry in the lower right corner is optional, as denoted by the square brackets. If it is 0, then all individuals die after the last age. If it is 1, then the last age class is an open ended interval, with individuals continuing to experience the transitions and mortality defined by **U**_*ω*_.^5^

The population (or cohort) structure at any time is given by the vector

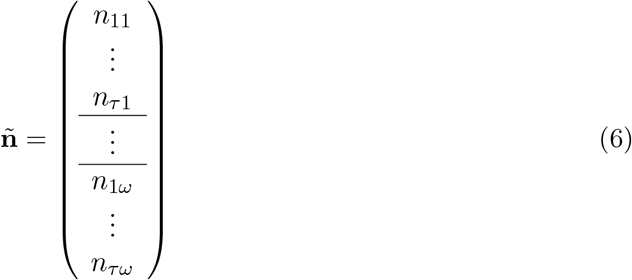

in which health stages are grouped with age classes. The multistate age*×*stage-classified transition matrix operates on a vector with this structure. To construct the transition matrix, define block-diagonal matrices, of dimension (*τω × τω*),

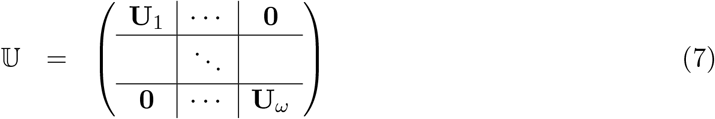

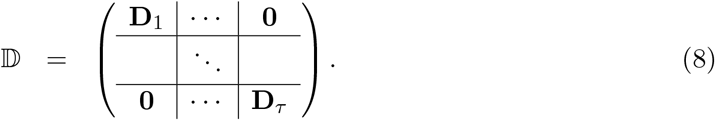

Then the block-structured matrix that forms the transient part of the age*×*stage-classified transition matrix is

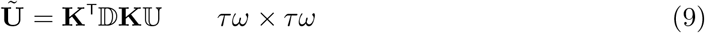

Here, the matrix **K** is a special permutation matrix known as the vec-permutation matrix (Henderson and Searle, 1981); see AppendixB for calculation.

The mortality matrix **M** is of dimension *α × τ*. The (*i, j*) entry of the matrix **M**_*x*_ is the probability of transition from the *j*th transient state to the *i*th absorbing state, conditional on age *x*. The matrix 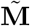 combining these matrices is

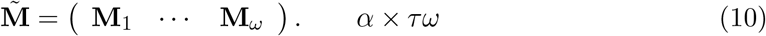

The transition matrix for the age-stage classified Markov chain is then

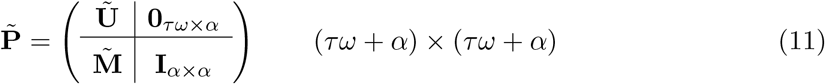

By this process, we have reduced the apparently complicated age-stage model to the same form as a simple age- or stage-classified absorbing Markov chain.

### 3.2 A model for colorectal cancer

As an example of a Markov chain model for health stages, we consider the model due to Wu et al. (2006) for colorectal cancer (CRC). Colorectal cancer is the second most common cancer in women, the third most common in men, and the fourth most common cause of death from cancer (Stewart and Wild, 2014). The development of CRC begins as abnormal but benign cell growths (adenomas) in the colon or rectum. It can then progress to cancerous cells within the intestine (adenocarcinoma) and eventually to metastatic cancer (Stewart and Wild, 2014). Wu et al. (2006) developed a Markov chain model for the progression of CRC as part of a program designed to study the costs and benefits of alternative screening procedures.

The stages of the disease and the transition structure among those stages are shown in Figure 1. In the early preclinical CRC stages (Dukes’ stages A and B) the cancer is restricted to within the colon. The late preclinical stage (Dukes’ stages C and D) involve metastasis to the lymph nodes or elsewhere in the body. The preclinical stages can eventually reach the point of requiring clinical intervention.

**Figure 1:**
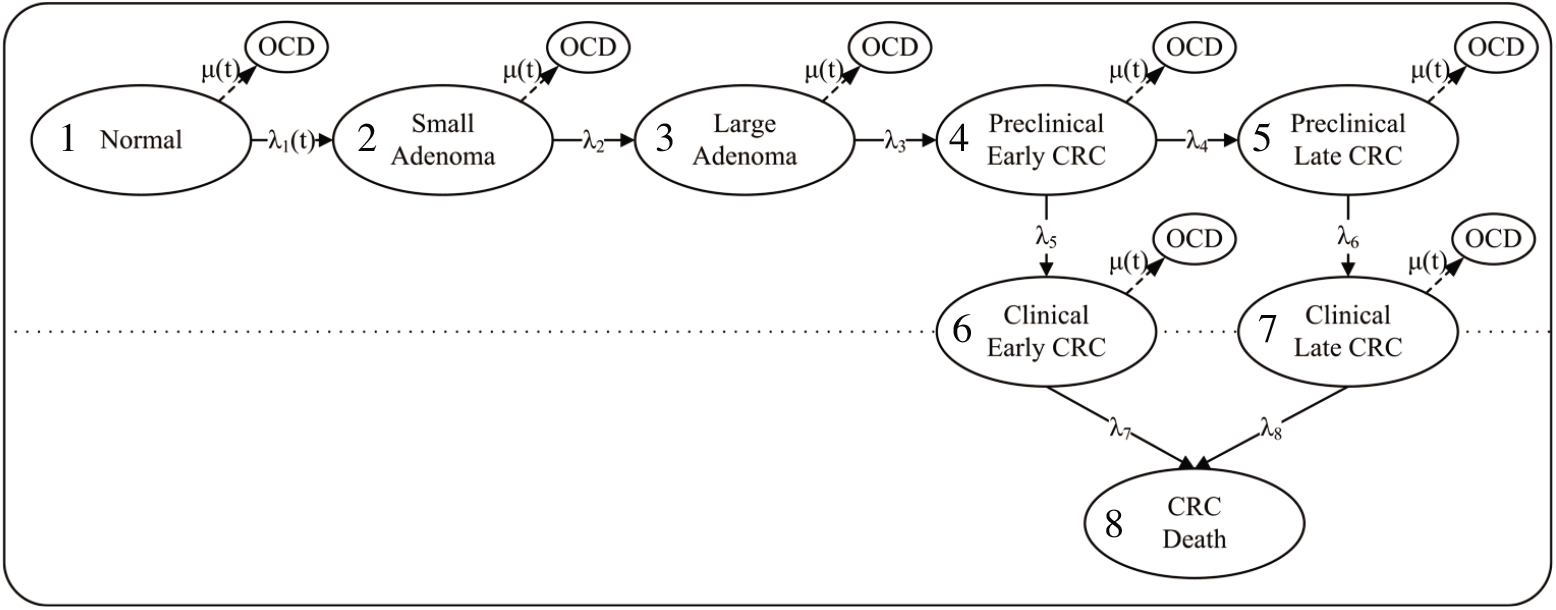
The transition structure for the stages of colorectal cancer (CRC). OCD death from causes other than CRC. Modified from Figure 1 of Wu et al. (2006) under the terms of a Creative Commons Attribution license.

The model of Wu et al. (2006) is a continuous-time Markov chain specified by rates *λ*_*i*_ for movement among stages and *µ*_*i*_ for mortality due to other causes of death. The transition rate *λ*_1_(*x*) from normal cells to small adenoma is the only age-dependent transition rate, described by Wu et al. (2006) as a Weibull function. The values listed in Table 1 of Wu et al. (2006), at five-year age intervals, are nearly linear starting at age 50, so for our purposes we interpolated them to one-year intervals and extrapolated them to age 100. The mortality rate *µ*_*i*_ due to causes other than colorectal cancer was obtained from the age-specific mortality, both sexes combined, for Taiwan in 2002 (Human Mortality Database, 2019).

For each age class *x* we constructed the intensity matrix **Q**_*x*_ following Figure 1, calculated the discrete skeleton of the model as **P**_*x*_ = exp **Q**_*x*_, and extracted the transient matrices **U**_*x*_ and the mortality matrices **M**_*x*_ from **P**_*x*_ as in equation (1). Because mortality due to CRC and to other causes is included in the **U**_*x*_, the age advancement matrices **D**_*j*_ contain ones on the subdiagonal and zeros elsewhere, as in (5). The multistate transient matrix **Ũ** and the multistate mortality matrix 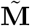 were constructed as in (9) and (10). The results of these and other calculations are contained in the Matlab code in the Supplemental Materials.

## 4 Markov chains with rewards

Healthy longevity is an occupancy time and our goal is to compute the statistics of the time spent in some specified set of health conditions. We confront a mathematical problem here. We are, in general, interested in occupancy not of a single state of the multistate Markov chain, but a set of states, defined by certain combinations of ages and health stages. Classical Markov chain theory provides all the moments of the occupancy time for any single state but not for sets of states (e.g., mildly or severely disabled at ages between 60 and 70). Thus classical theory could provide the statistics of longevity for, e.g., early preclinical CRC at age 60, but not for more interesting combinations such as cancer-free longevity between 60 and 70.

We solve this problem^6^ by using Markov chains with rewards (MCWRs); these permit a stochastic analysis of healthy longevity for arbitrary combinations of ages and health stages. We will see below what this implies in practice for the CRC model, but first we turn to the basic methodology of MCWRs.

Life is a Markov chain; health is a reward. An individual in a Markov chain makes transitions among states. A Markov chain with rewards (MCWR) imagines that the individual collects a “reward” *r*_*ij*_ when making the transition from state *j* to state *i*. These rewards accumulate over the lifetime of the individual. The lifetime accumulation of the reward might measure lifetime occupancy of a health stage, lifetime accumulated quality of life or lifetime numbers of entries to a hospital. The analysis provides an extremely flexible definition of healthy longevity.

MCWR methods were introduced by Howard (1960) as the basis for stochastic dynamic programming (e.g., Puterman, 1994; Sheskin, 2010). Extensions of the theory to random rewards and demographic models were introduced by Caswell (2011) and have been extended with applications to lifetime reproductive output (Caswell, 2011; van Daalen and Caswell, 2015; van Daalen and Caswell, 2017), evolutionary biodemography (van Daalen and Caswell, 2020), income and expenditures (Caswell and Kluge, 2015), and healthy longevity in prevalence-based models (Caswell and Zarulli, 2018; Owoeye et al., 2020).

The reward *r*_*ij*_ is a random variable and is characterised by its moments. Reward matrices can be estimated in many ways, depending on the nature of the reward (van Daalen and Caswell, 2017). For the incidence-based health model, we require rewards for transitions between all age-stage combinations.

Since the elements of the reward matrices correspond to the entries of the transition matrix, we write the reward matrices with a block structure corresponding to that of the transition matrix **P**. We define 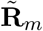 as the reward matrix containing the *m*th moments of the rewards. That is,

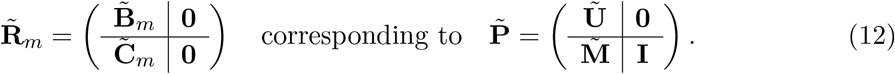

That is, the entries of 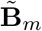 are the *m*th moments of the rewards associated with the corresponding transitions in **Ũ**. The entries of 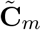 are the *m*th moments of the rewards associated with the transitions, which appear in 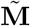, from transient to absorbing states.

### Lifetime rewards

Rewards are accumulated from any starting state until absorption. No rewards are accumulated after death (i.e., there is no healthy longevity after death; this seems like a safe assumption). The moments and the statistics (mean, variance, coefficient of variation, skewness, etc.) of the lifetime accumulation are calculated from the Markov transition matrix 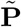 and the reward matrices 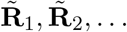,.

The formulae^7^ for the accumulated rewards are given by van Daalen and Caswell (2017, Theorem 1). The first three moments are, in our notation,

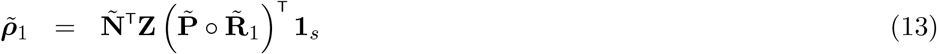

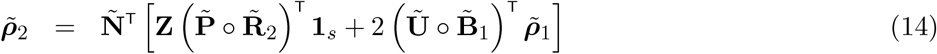

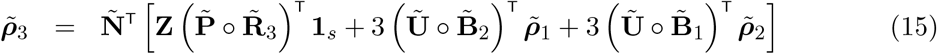

where **Ñ** is the fundamental matrix,

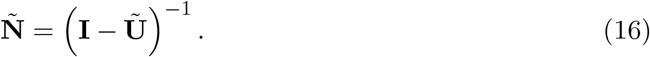

The (*i, j*) entry of **Ñ** is the mean time spent in state *i*, before eventual absorption, conditional on starting in state *j*, with *i* and *j* ranging over all the age*×*stage combinations. The matrix **Z** slices off the rows and columns of 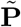 and 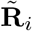 corresponding to the absorbing states,

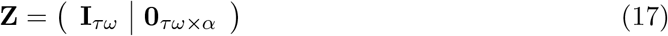

The matrix 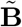 is the submatrix of 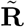 corresponding to the transient states, as in equation (12). The vectors 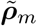 are all of dimension *τω ×* 1. See (2017) for all the moments.

The first three moments suffice to calculate the mean, variance, and skewness of healthy longevity:

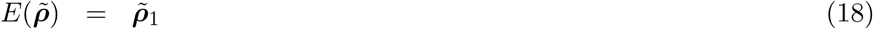

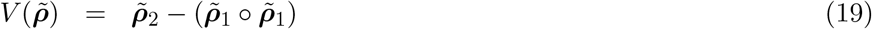

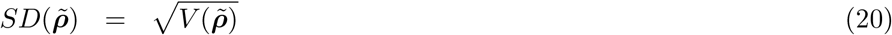

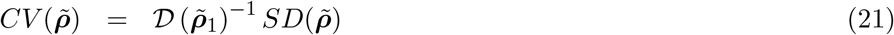

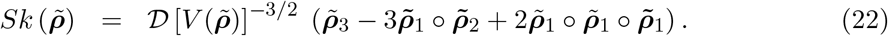

The vector 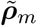 contains the *m*th moments of healthy longevity for all combinations of initial age and health stage. To obtain the moments of healthy longevity as a function of age for a specified initial stage *i*, we calculate

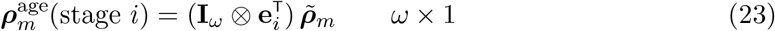

where **e**_*i*_ is the *i*th unit vector of length *τ*. Similarly, the moments as a function of stage for a specified initial age *x* are given by

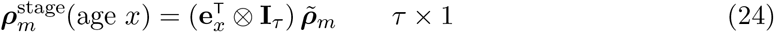

where **e**_*x*_ is the *x*th unit vector of length *ω*.

Health and healthy longevity are protean concepts with many definitions. Each has its own reward structure, and in the next section we show how to calculate the components 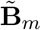 and 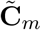 of the reward matrices 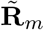 in equation (12).

## 5 Health reward structures and defining healthy longevity

We consider four ways to define healthy longevity.

1. **Occupancy of health states**. The most basic definition of healthy longevity is the length of life spent in some specified health condition. The health condition of interest may be defined by any combinations of ages and health stages.
2. **Transitions among health states**. Rather than the time spent in a specified health condition, one might define healthy longevity in terms of the lifetime number of transitions among specified health stages at particular ages.
3. **The “value” of occupancy of health states**. Healthy longevity might be something more nuanced than simply the occupancy of particular sets of stages. Instead we might assign a value (e.g., the quality of life) to each stage.
4. **The “value” of transitions among states**. As with occupancy, it may be useful to assign values to transitions among particular health stages at particular ages.

### 5.1 Healthy longevity as occupancy

The goal is to calculate the lifetime occupancy of a health condition of interest. The health condition might be simple (healthy vs. not) or more complicated (early stage illness with long-term institutional care…). The lifetime might include all ages, or some specified set of ages of interest. To create the reward matrices, we begin with an array; let us call it a health occupancy matrix **H**. The rows of **H** correspond to health stages and the columns to ages. For the colorectal cancer model of Figure 1, **H** has the form

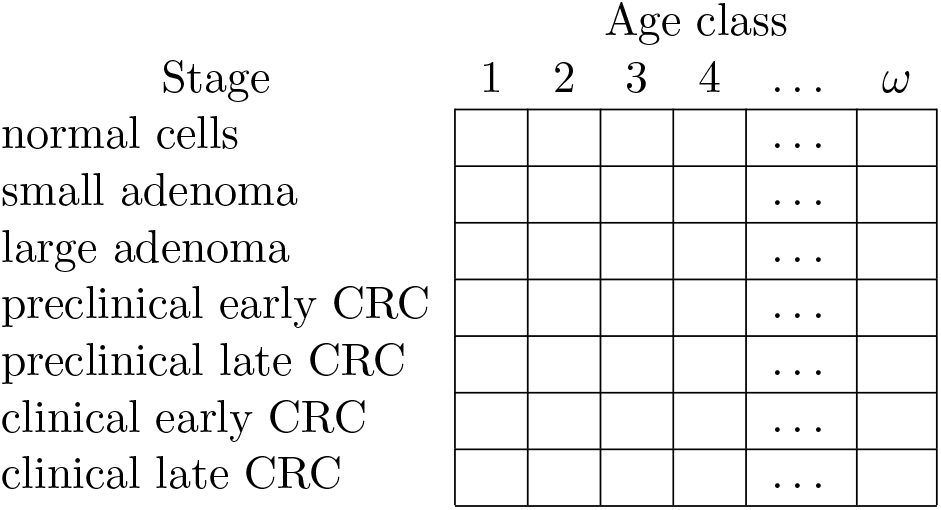

Define a set 𝒮 of age-stage combinations that will be considered healthy longevity:^8^

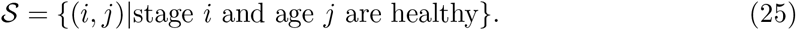

Enter 1 in the cells of **H** corresponding to the elements of 𝒮 and 0 elsewhere; i.e.,

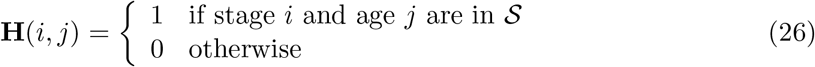

#### 5.1.1 Examples in the colorectal cancer model

Here are four examples that will be explored below in the analysis of the CRC model.

a. “Normal cell” longevity. This is the time spent in stage 1 over a lifetime, without any abnormal or cancerous cells; **H** is shown in Figure 2(a).
b. Cancer-free longevity. This is the spent in any of the cancer-free stages (1–3) over a lifetime. In this case *S* contains all entries of rows 1–3 of **H**, as shown in Figure 2(b).
c. Early cancer. Suppose that healthy longevity is defined as the time spent in any of the clinical cancer stages (6 and 7) prior to a specified age. For example, in the United States, individuals become eligible for Medicare health insurance at age 65. The implications of clinical CRC before and after age 65 can be very different. The health occupancy matrix **H** is shown in Figure 2(c) for the hypothetical case of longevity in clinical cancer stages prior to age class 4.
d. Late cancer. As in (c), but counting time spent in any of the clinical cancer stages after some specified age, as shown in Figure 2(d)

**Figure 2:**
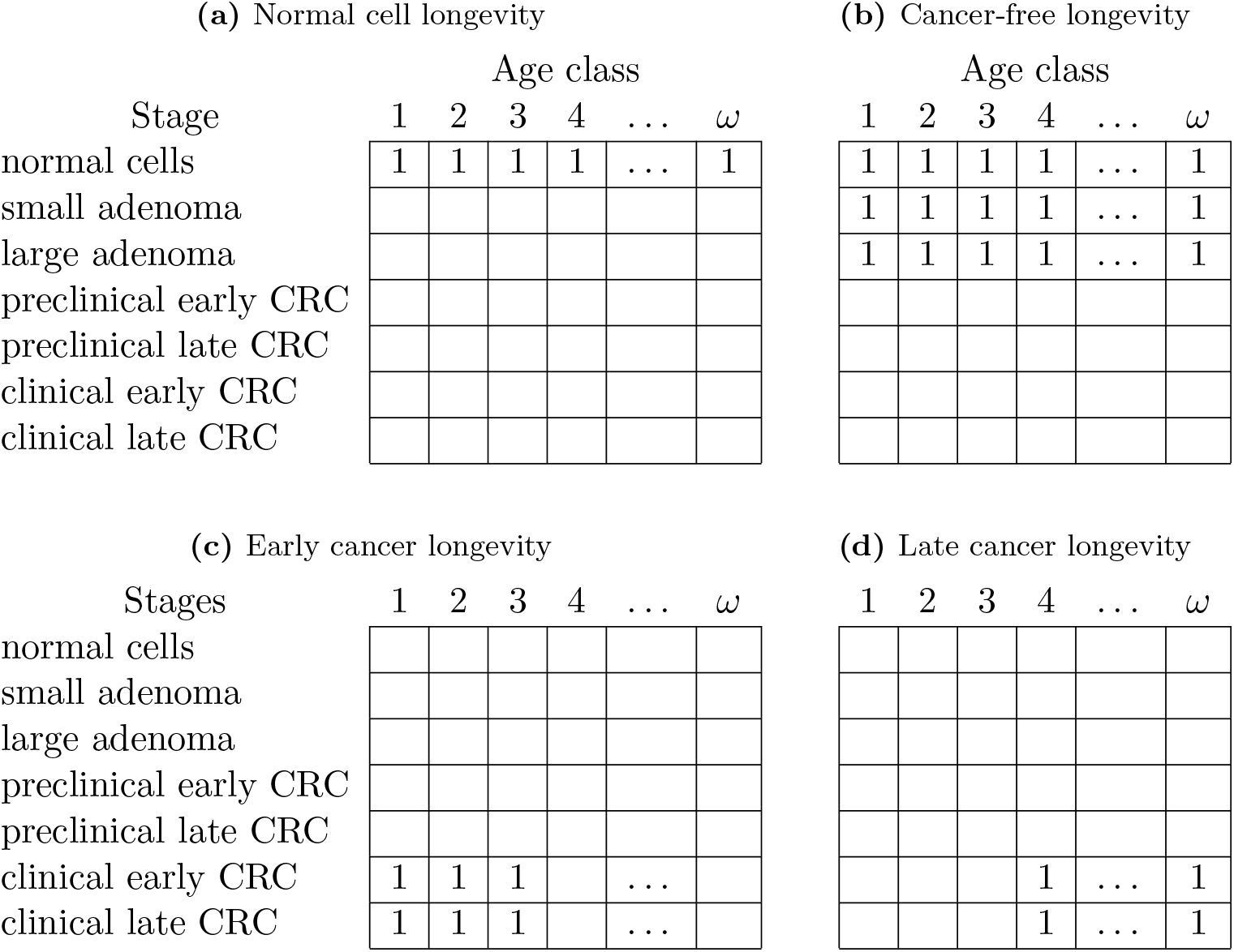
Examples of the health occupancy matrix **H** for specified definitions of longevity for the colorectal cancer model. Empty cells are understood to contain zeros. (a) Normal cell longevity, measured by time spent in the normal cell stage at any age. (b) Cancer-free longevity, measured by the time spent, at any age, in any of the stages 1–3. (c) Early cancer longevity, measured by the time spent in either of the clinical CRC stages prior to some age, here taken as age class 4. (d) Late cancer longevity, at age 4 or older.

The number of possible definitions of healthy longevity is truly astronomical. The colorectal cancer model contains seven health stages and 50 ages, for a total of 350 age-stage combinations. Each may receive a 0 or a 1 in **H**, for a total of 2^350^ ≈ 2 × 10^105^ possible definitions of healthy longevity. It is said that the number of stars in the visible universe is only (ha!) on the order of 10^24^. Even the number of atoms is only about 10^78^ (https://www.universetoday.com/). Any of these definitions could be analysed using **H**, making the approach extremely flexible. It is hard to imagine why you would want to know about longevity in, say, odd-numbered health states in prime-numbered ages, but if you want to, you can. Note that total longevity, independent of health status, is given by inserting a 1 in every entry of **H**.

### The reward indicator vector

The next step is to create an occupancy indicator vector by applying the vec operator to **H** (stacking the columns on top of each other);

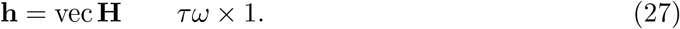

This 0-1 vector inherits the block structure of the vector **ñ** in (6) and the age-stage matrices **Ũ**, etc. Also define a vector ¬**h** that contains the logical complement of the entries of **h**; i.e.,

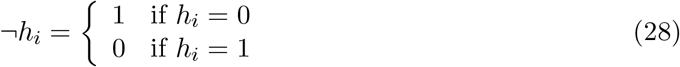

### Partial rewards for partial occupancy

If an individual spends the entire interval within the set 𝒮 of age-stage combinations, the result is an unequivocal occupancy of one time interval. But what about individuals that leave the set during the interval (by death, or by moving to an age-stage combination not in 𝒮)? What about individuals that enter the set during the interval (from age-stage combinations not in 𝒮)? (This issue was suggested by Daniel Schneider and Mikko Myrskylä; see also Sonnenberg and Wong 1993) Demographers solve this problem when calculating mortality rates from counts of individuals and numbers of deaths by defining a fraction of year lived by those who die. The fraction is often denoted by *a*_*x*_ in life table presentations; *a*_*x*_ is often taken to be 0.5, indicating that individuals that die are alive, on average, for one-half of an age interval. An exception is usually made for the first year of life, when infant mortality shifts the fractional occupancy to something smaller, perhaps 0.1 (Chiang, 1968; Preston et al., 2000).

We apply the same logic to partial occupancy here. We define *a* as the fractional occupancy experienced by an individual that enters or leaves the set 𝒮 during the interval, and set 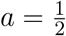.

### The reward matrices for longevity

We need the moment matrices 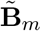 and 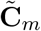 that appear in (12). The calculation of 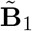 must account for three kinds of occupancy:

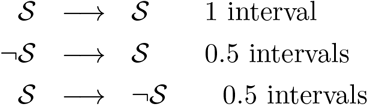

Recalling that **h** indicates the states in 𝒮 and ¬**h** the states in ¬ 𝒮, the matrix is

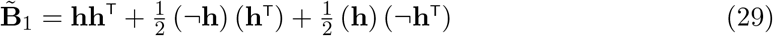

The first term gives a unit reward to an individual starting and ending the interval in any of the states within 𝒮. The second term gives a reward of 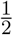 to an individual that starts the interval within 𝒮 but finishes the interval outside 𝒮. The third term gives a reward of 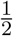 to an individual that starts the interval outside of 𝒮 but finishes the interval within 𝒮. Because **h** + ¬**h** = **1**, the result (29) reduces to the (remarkably simple)

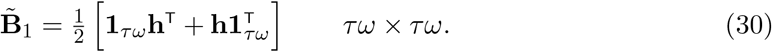

Transitions into absorbing states from a state in 𝒮 earn a fractional occupancy reward; the component 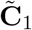 is given by

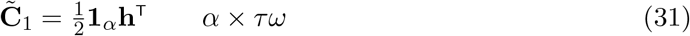

The matrix of first moments of rewards, 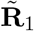, is assembled from 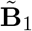 and 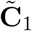 as in (12). Because occupancy is a fixed reward (sensu Caswell 2011; van Daalen and Caswell 2017) the second and third moment matrices are

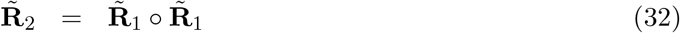

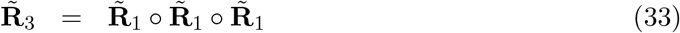

and, in general,

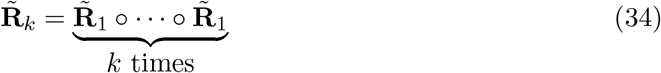

This provides all the pieces needed to compute the moments of healthy longevity using equations (13)–(15) and associated statistics from (18)–(22).

### 5.2 Healthy longevity as transitions

Instead of counting the time occupying a set of age-stage combinations, we may measure healthy longevity by counting transitions among health stages. For example, transitions into and out of care facilities have been studied in the context of cognitive impairment (e.g., Tyas et al., 2007; Coley et al., 2015), nursing home use (Hurd et al., 2014), and hospital admissions (Ieva et al., 2017). It may be important to count transitions at particular ages, and different transitions may be of interest at different ages.

Suppose that, at age *x*, there are *N* transitions of interest. Define the set of these transitions as

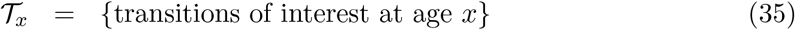

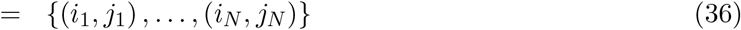

where (*i, j*) denotes the transition *j* → *i*. The set 𝒯_*x*_ may include transitions into absorbing states as well as transitions among transient states.

For example, the CRC model in Figure 1 contains two transitions into clinical (metastatic) disease ((6, 4) and (7, 5)). To count the number of entries^9^ by an individual into clinical CRC care over a lifetime or some part of a lifetime, the set 𝒯 would be

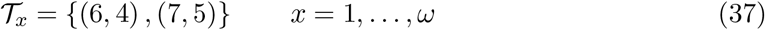

At each age *x*, create matrices 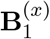 and 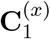 that indicate the transitions contained in 𝒯_*x*_

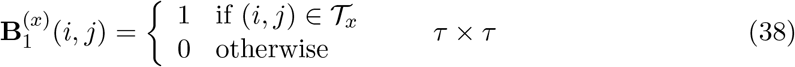

where *i* and *j* denote transient states. Transitions into absorbing states are counted by

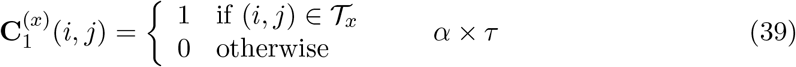

where *i* is an absorbing state and *j* is a transient state.

In the case of the CRC model, counting entries into clinical CRC would yield, for any age *x*,

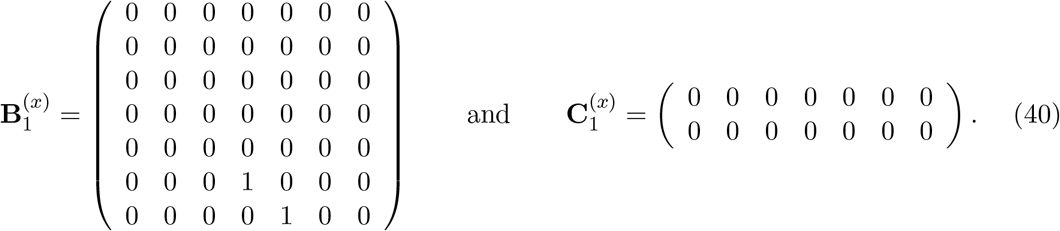

The age-stage matrix 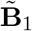 of first moments is obtained using the same vec-permutation construction that produced **Ũ**,

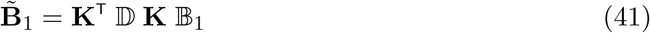

where

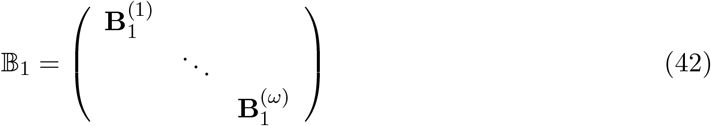

and 𝔻 is as defined in (8). The matrix 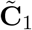 is

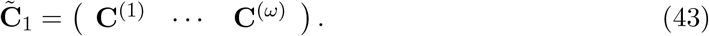

The reward matrix 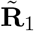 is assembled as in (12). Because transitions, like occupancy, are fixed rewards, the higher moments are given in equation (34).

Occupancy and transition calculations can be combined to compute the number and duration of ‘episodes.’ Dudel and Myrskylä (2020) used MCWR to calculate the expected number of transitions into a state and the expected time occupying the state. The occupancy time divided by the number of entries is the mean length of an episode; they applied the calculations to a model for liver cirrhosis and for disability in the elderly. Our approach would permit easy generalisation of their calculations to a full age×stage-classified multistate model, with episodes defined in terms of sets of health stages and age classes as desired.

### 5.3 Assigning a value to occupancy

Suppose that values are assigned to the time spent in particular age-stage combinations. The values might measure a benefit (e.g., quality of life, or the chance of labor force participation) or a cost (e.g., disability, or expense of medical treatment). This value is a random variable characterised by a set of moments. Let *v*(*i, j*) denote the value attached to one unit of time spent in health stage *i* at age *j*. Create a set of arrays **V**_*m*_, with the same structure as **H**, where **V**_*m*_(*i, j*) is the *m*th moment of the value of a unit of time in stage *i* at age *j*.

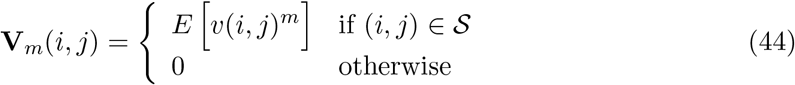

Define moment vectors in the same way as the occupancy indicator vector **h**.,

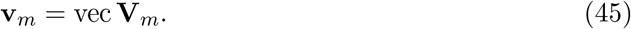

Let us assume that partial occupancy for 1/2 a time unit has a value 1/2 of the random value *v*(*i, j*). Then

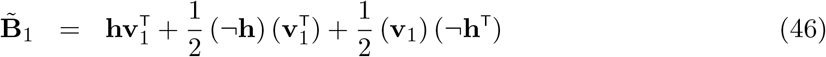

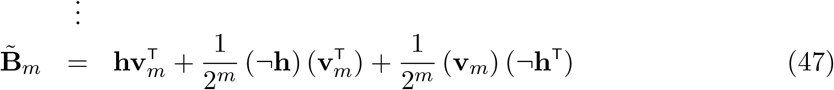

Transitions into absorbing states accumulate value only for the fraction of the interval in the state of origin, so that

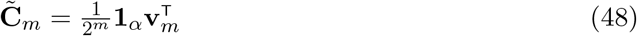

The reward matrices 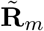 are assembled from 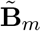 and 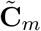 as in equation (12). Rewards based on the value of occupancy need not be fixed, so 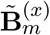 and 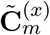 cannot be constructed from 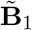 and 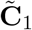, but must be constructed from data or assumptions about the moments of the values. Note that if the value of a unit of occupancy is simply the occupancy itself, so that **v** = **h**, then (46) reduces to (29), as it should.

Other models are available for allocating value to partial occupancy. For example, value might take the form of a single packet or event arriving at a time uniformly distributed over the interval. Then half a unit of occupancy would mean a probability 0.5 of receiving one packet and a probably 0.5 of receiving none. Or, perhaps value is a count of things that arrive at random during the interval as a Poisson process, and the value of half a unit is the result of that Poisson process or one half interval. These options are explored in Appendix C.

### 5.4 Assigning a value to transitions

Transitions from one health state to another often incur costs or benefits (e.g., entering a hospital might have special admission costs, or different insurance coverage). Those costs and benefits will generally vary with age, and we would like to assign values to health stage transitions as a function of age.

Transitions are, as in Section 5.2, defined by an origin stage, a destination stage, and an age class. The set of transitions of interest at age *x* is 𝒯_*x*_. Let *v*(*i, j, x*) be the value attached to a transition *j* → *i* at age *x*. The matrices **B**^*(x)*^ and **C**^*(x)*^ defined in equations (38) and (39), now become

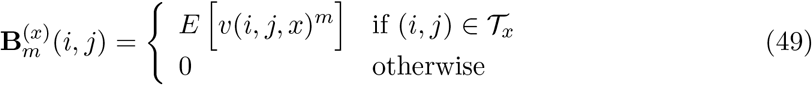

if *i* and *j* are transient states, and

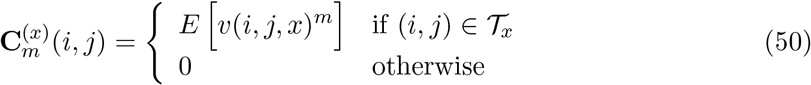

when *i* is an absorbing state and *j* is a transient state.

Given these age-specific matrices, the block-structured matrices 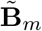 and 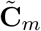 are calculated using the vec-permutation matrix as in (41) and (43). The reward matrices are then given by equation (12).

Unlike the rewards for counting transitions, rewards based on the value of transitions are not fixed (i.e., not every individual making the transition receives the same reward) and hence the moment matrices must be calculated from the moments of the distribution of *v*(*i, j, x*). Of course, if, every transition in 𝒯_*k*_ is given a fixed value of 1, the value of transitions reduces to the count of transitions, as in Section 5.2.

A particularly important transition is the transition to death. It happens only once, and eventually happens to everyone, so counting the number of transitions may be of limited interest. But the death of an individual is associated with a value (a cost, in this case): the years of life lost (YLL) due to that death. Vaupel and Canudas Romo (2003) introduced this concept in the context of changes in mortality and denoted its expectation by *e*^†^. But the years of life lost due a death is a random variable, and by treating it as a value attached to the transition to death, we are now in a position to calculate all the moments of YLL. Caswell and Zarulli (2018) showed how to do this for a prevalence-based model, in which individuals are classified only by age. In such models, YLL is one component of disability-adjusted life years (DALYs), a primary component of the Global Burden of Disease Study (e.g., Devleesschauwer et al., 2014; GBD 2015 DALY and HALE Collaborators, 2016). With an incidence-based model, it is possible to compute all the moments of life lost, beginning in any initial age and health stage, and accounting for the dependence of remaining longevity on both age and health stage; see Section 6.2 below.

The first three moments of remaining longevity are given by the entries of the vectors

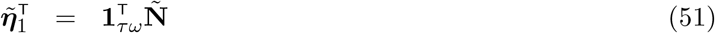

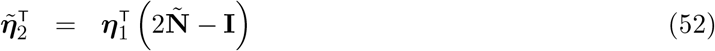

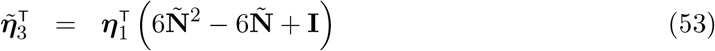

where **Ñ** is the fundamental matrix given in (16) and 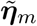 is the (block-structured) vector of the *m*th moments of longevity (Iosifescu, 1980; Caswell, 2013).

Suppose, as is true in the example in Section 6, that we distinguish two causes of death, and we are interested in years of life lost to cause 1. In the transition matrix (11), the probabilities of transition to death from cause 1 appear in row 1 of the mortality matrix 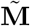.

Then using the formulae in equations (49) and (50) we obtain

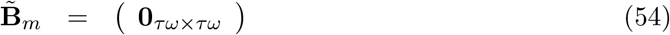

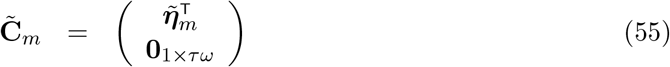

Following the construction in equation (12) gives the reward matrices

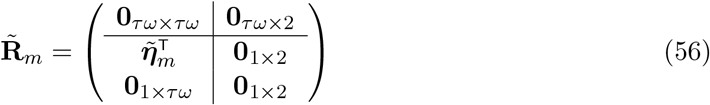

The age×stage-structured vector 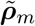 computed from **Ũ**, 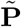, and the reward matrices contains the *m*th moments of life lost, due to death from cause 1, of every age-stage combination.

## 6 An example: healthy longevity in a colorectal cancer model

As an example, we present some results for healthy longevity, in terms of occupancy and of transitions, for the CRC model of Wu et al. (2006).

### 6.1 Healthy longevity and CRC: occupancy

The model contains 7 transient (living) stages, and 50 age classes (50–100 years). We will show results for the four examples of healthy longevity defined in Section 5.1: normal cell longevity, cancer-free longevity, early cancer longevity, and late cancer longevity. Early and late are defined here relative to age 65. Figures 3–6 compare the remaining healthy longevity, as a function of age, for individuals starting in two initial health stages: normal cells and large adenoma. Of course, an individual 65 or older who is in the normal or large adenoma stage cannot develop early cancer, and thus will have zero occupancy in that category.

**Figure 3:**
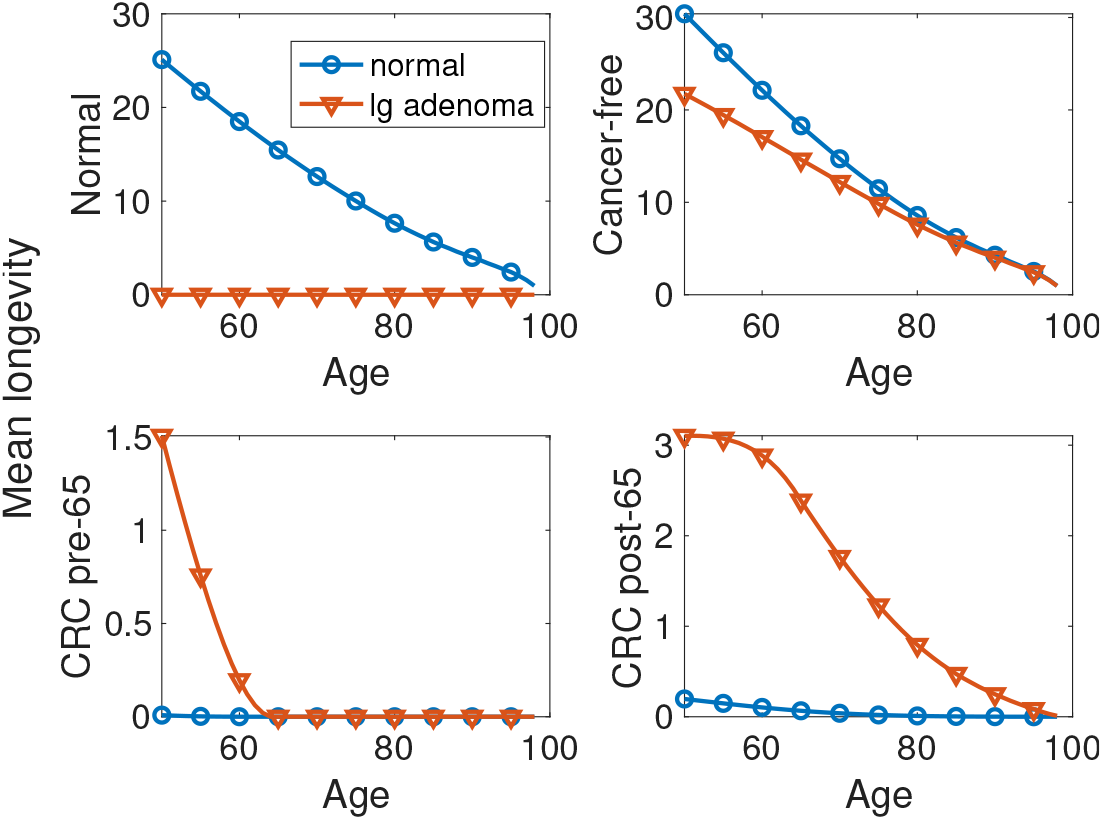
Mean remaining longevity for an individual starting in the stages normal cells and large adenoma, as a function of age. Longevity is defined as occupancy time in the sets of stages: normal cells, cancer-free, early CRC and late CRC (relative to age 65).

**Figure 4:**
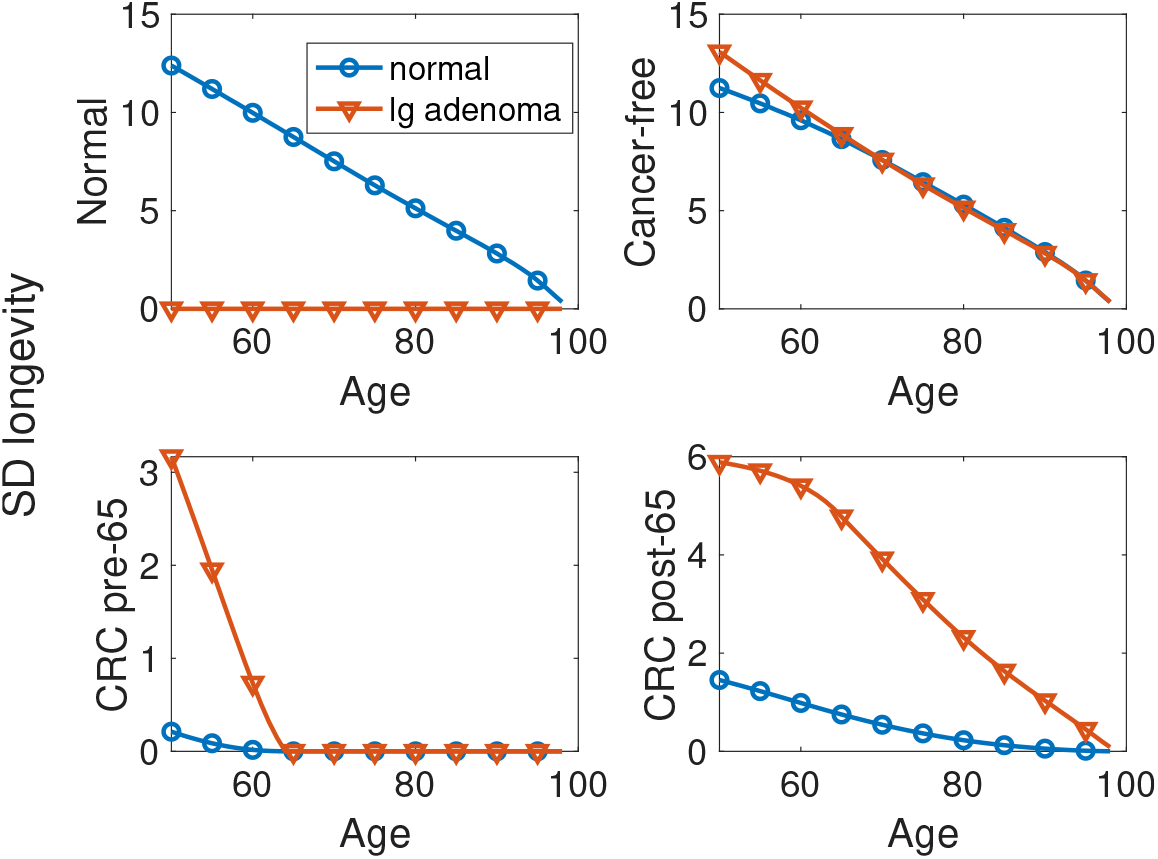
Standard deviation of remaining longevity for an individual starting in the stages normal cells and large adenoma, as a function of age. Longevity is defined as occupancy time in the sets of stages: normal cells, cancer-free, early CRC and late CRC (relative to age 65).

The mean healthy longevity (health expectancy) shows the expected pattern of decline with age (Figure 3). Individuals with large adenoma have about a 10 year disadvantage in cancer-free longevity at age 50, compared with normal cell individuals. The difference declines with age and becomes negligible after about age 80. Individuals with large adenomas have a dramatically increased risk of clinical cancer, either before or after age 65. The most dramatic difference is the time spent with CRC before and after the age of 65. The presence of large adenoma greatly increases the mean lifetime spent with colorectal cancer, both before and after age 65.

Moving on to measures of inter-individual variability, we see that the standard deviation of healthy longevity is large (Figure 4). At age 50, for example, cancer-free longevity has a standard deviation of 10–15 years. The standard deviation of longevity with CRC, either pre- or post-65, is increased many-fold in individuals with large adenoma, compared to those with only normal cells.

Figure 5 shows the coefficient of variation (CV), which scales the standard deviation relative to the mean. Healthy longevity in the normal and the cancer-free conditions is reasonably predictable by this measure, with CV less than 1. In contrast, the time spent with clinical CRC for individuals in the normal cell state is very uncertain, with a CV much greater than 1. This is especially true for older individuals.

**Figure 5:**
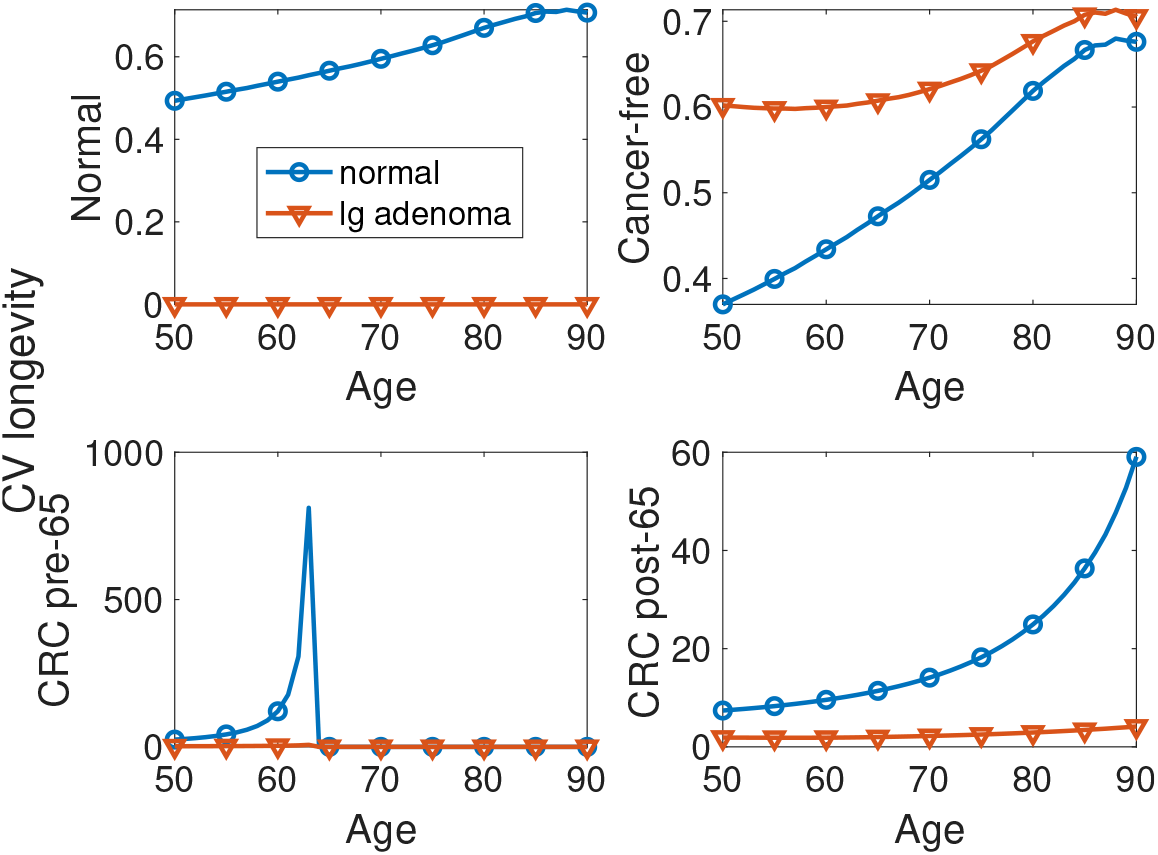
Coefficient of variation (CV) of remaining longevity for an individual starting in the stages normal cells and large adenoma, as a function of age. Longevity is defined as occupancy time in the sets of stages: normal cells, cancer-free, early CRC and late CRC (relative to age 65).

The skewness of healthy longevity in these conditions emphasises this uncertainty (Figure 6). The skewness of longevity in the normal cell and the cancer-free states is small, negative up to about age 70 and positive after that. But longevity with early or late clinical CRC is extremely positively skewed; some small fraction of individuals are expected to spend a much longer time in these CRC conditions than will typical individuals, and a focus on mean longevity seriously underestimates these extreme cases.

**Figure 6:**
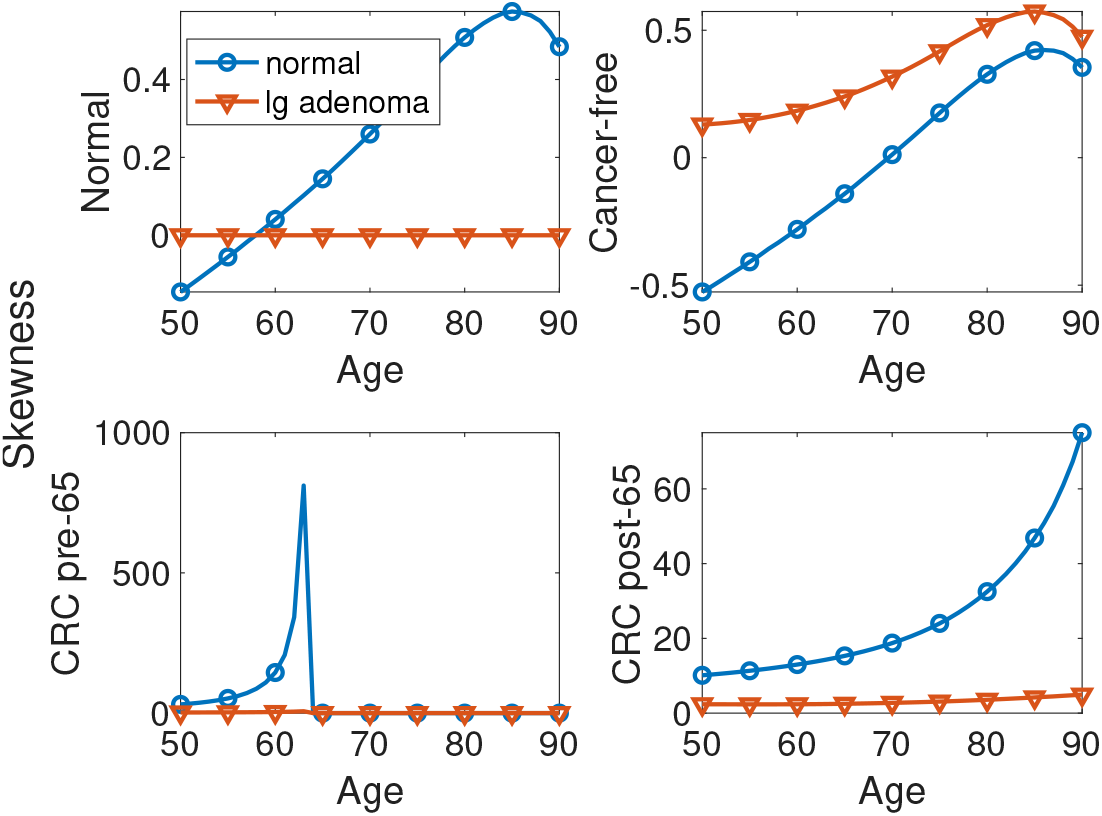
Skewness of remaining longevity for an individual starting in the stages normal cells and large adenoma, as a function of age. Longevity is defined as occupancy time in the sets of stages: normal cells, cancer-free, early CRC and late CRC (relative to age 65).

An intuitive feel for this uncertainty is provided by the following calculation. Imagine picking, at random, two individuals identical in age class and health stage. How similar will their eventual healthy longevities be? What is the chance that those longevities will differ by a factor of 2? or a factor of 10? The higher the probability of a *k*-fold difference, the more uncertain the healthy longevity. In the context of bioassay studies, random pairs of replicate measurements are compared as part of quality control. Reed et al. (2002) derived the probability that two samples (two individuals in our case) will differ by a factor of *k*, as a function of the CV of the measurement (healthy longevity in our case):

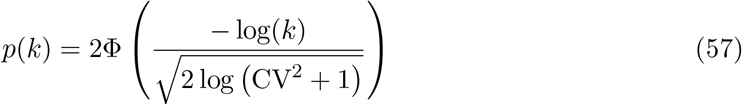

where Φ(·) is the cumulative standard normal distribution. Their analysis is based on a lognormal approximation; the lognormal is a flexible distribution that should provide some information for us.

Figure 7 shows the probability of a 2-fold difference between randomly selected individuals in each of the measures of healthy longevity. The probabilities are high, especially for the longevity in the early and late clinical CRC conditions. The late CRC longevity of two randomly selected individuals, aged 50 with normal cells, will differ by a factor of at least 2 more than 80% of the time. We offer this example as indicative of the type of uncertainty that can exist in healthy longevity; further development of such analyses would be valuable.

**Figure 7:**
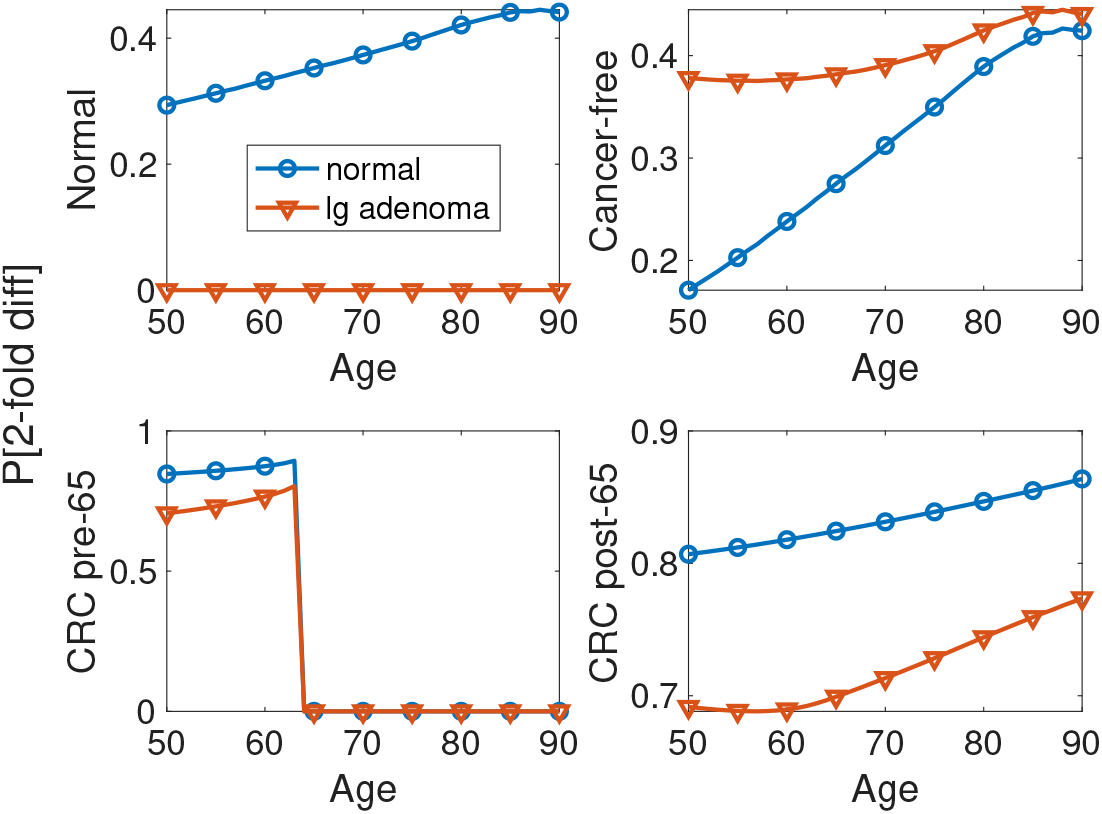
Probability of at least a 2-fold difference in remaining longevity between two randomly selected individuals. Longevity is defined as occupancy time in the sets of states: normal cells, cancer-free, early CRC, and late CRC.

### 6.2 Healthy longevity and CRC: transitions

#### 6.2.1 The transition to clinical cancer

The CRC model contains two stages (6 and 7) with clinical cancer. Transitions into these stages would be particularly significant for both individual health and for the kind of care and treatment required. So, we compute the statistic of the lifetime number of these transitions (4 → 6 and 5 → 7).

The matrices 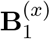 and 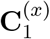 are given in equation (40). Since these transitions are irreversible, the number of entries into the clinical CRC stages is either 0 or 1, and the expected number of transitions is just the probability of making the transition. Thus the mean lifetime number of transitions is also the probability that an individual at a specified age will eventually develop clinical CRC, by either of the two possible pathways.

Figure 8 shows the expected number of transitions for an individual of specified starting age and health stage. The lifetime probability of developing clinical CRC is lowest for individuals in the normal cell stage, and highest for individuals with late preclinical CRC, and, not surprisingly, declines with age.

**Figure 8:**
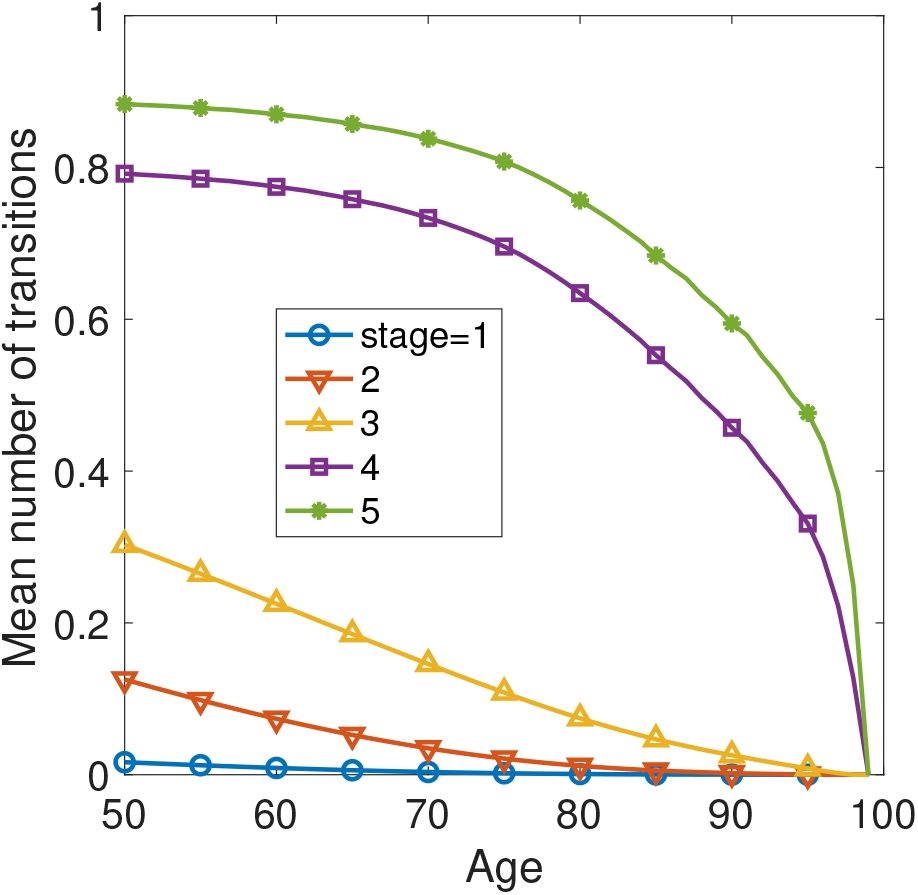
The expected number of transitions into stages 6 and 7 (clinical, metastatic CRC) as a function of individual age and health stage. Because stages 6 and 7 can be entered only once, this is also the probability of the transition into the stage.

#### 6.2.2 Years of life lost to CRC

The CRC model includes two causes of death: death due to cancer and death due to all other causes. Death due to cancer appears as a transition from stages 6 and 7 into the first absorbing state. As an example of the calculations in Section 5.4, we calculate the vectors of the moments of remaining longevity according to equations (51)–(53), and the reward matrices according to (56).^10^

Some results are shown in Figure 9. The statistics of YLL are dependent on age and strongly dependent on health stage. This example makes it clear that age alone does not provide reliable information on years of life lost due to CRC. The uncertainty in YLL, as measured by the standard deviation, increases when considering individuals with more advanced disease. When scaled relative to the mean, the uncertainty is enormous in the early stages (normal cells and adenomas). The years of life lost are positively skewed, especially at early health stages, indicative of a strong disparity between a few individuals losing many years and most individuals losing none or a few.

**Figure 9:**
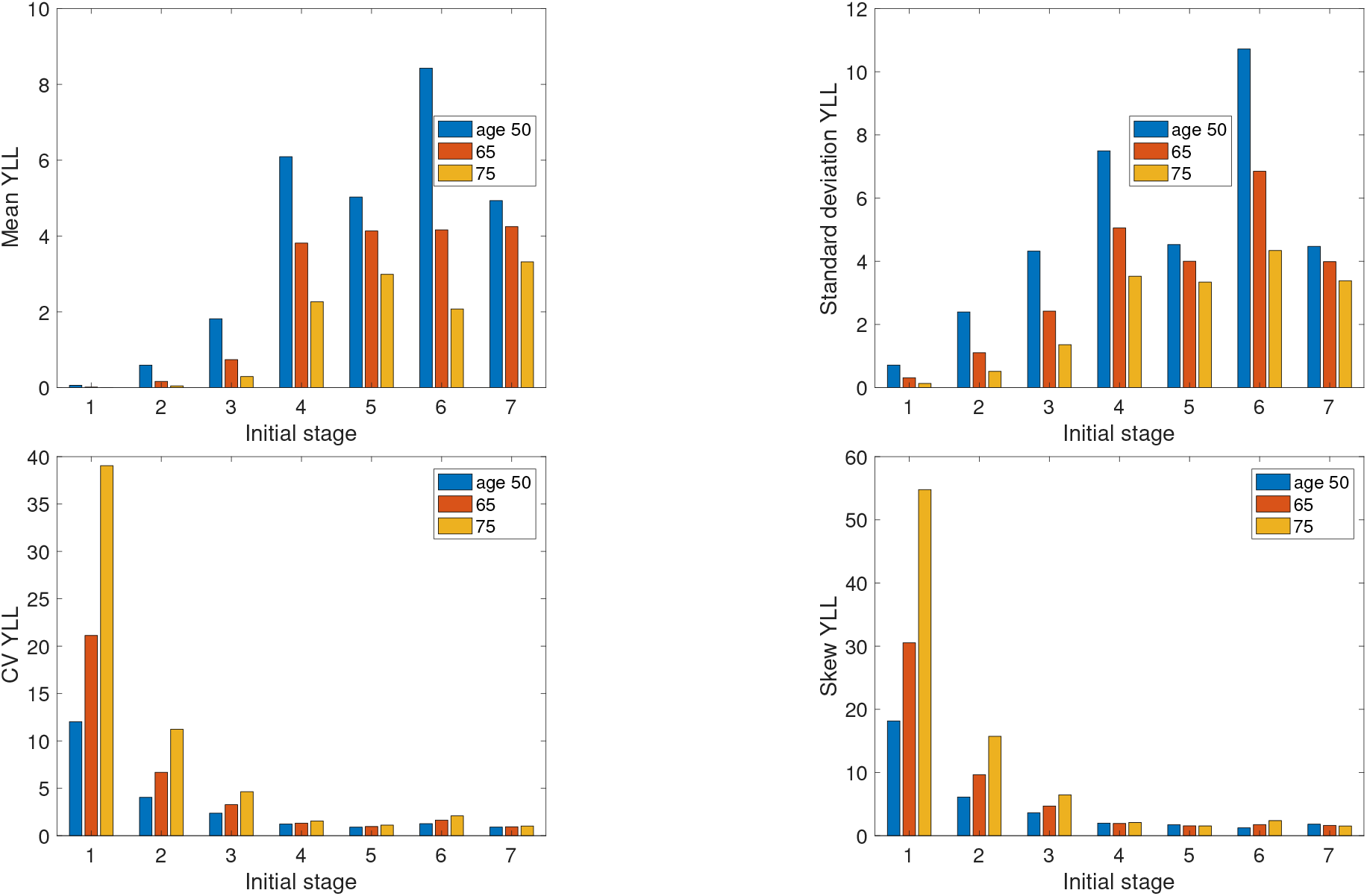
The mean, standard deviation, coefficient of variation, and skewness of years of life lost (YLL) to colorectal cancer for individuals of each different health stage at ages 50, 65, and 75 years. Stage numbers as defined in Figure 1

## 7 A protocol for calculating healthy longevity

Our results can be summarised in a step-by-step protocol for calculations of healthy longevity.

1. Start with the transition matrices **U**_*x*_ (*x* = 1,…, *ω*), the age advancement matrices **D**_*j*_ (*j* = 1,…, *s*), and the mortality matrices **M**_*i*_ (*i* = 1,…, *ω*).
2. Construct the block-structured age×stage-classified matrices **Ũ**, 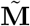, and 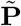 following equations (9), (10), and (11).
3. To calculate healthy longevity…
  … as lifetime occupancy of health stages, go to 4
  … as lifetime number of transitions among health stages, go to 5
  … as the value of lifetime occupancy of health stages, go to 6
  … as the value of lifetime transitions among health stages, go to 7
4. Healthy longevity as occupancy:
  a. Create the array **H** in Figure 2. Insert 1 in the ages and health stages of interest.
  b. Create the reward vector **h** by applying the vec operator to **H**, as in (27).
  c. Create the matrices 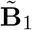 and 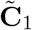 following (30) and (31).
  d. Create the matrix of occupancy rewards 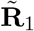 from 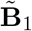 and 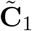 using equation (12).
  e. Create the higher moment reward matrices 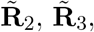 using (34).
  f. Go to 8
5. Healthy longevity as the number of transitions:
  a. For each age *x* of interest, specify the set 𝒯_*x*_ of transitions to count.
  b. Construct the indicator matrices 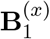 and 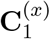 using (38) and (39).
  c. Construct the block-structured matrices 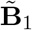 and 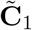 using (41) and (43).
  d. Assemble the matrix 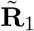 as in (12).
  e. Create the higher moment reward matrices 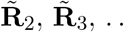… using (34) (f)
  f. Go to 8
6. Healthy longevity as the value of occupancy:
  a. Assign values to occupancy of each age-stage combination in matrices **V**_*m*_, for each moment *m* of interest, according to (44).
  b. Create the moment vectors **v**_*m*_ by applying the vec operator to **V**_*m*_, as in (45).
  c. Create the block-structured matrices 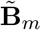 and 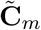 as in (47) and (48).
  d. Create the block-structured reward matrices 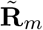 as in (12)
  e. Go to 8
7. Healthy longevity as the value of transitions:
  a. For each age *x*, obtain the moments of the value of each transition in 𝒯_*x*_.
  b. For each age *x* and each moment *m*, create the matrices 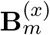 and 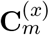 according to (49) and (50).
  c. Construct the block-structured matrices 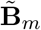 and 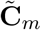 using (41) and (43) and assemble the matrices 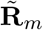 as in (12).
  d. Go to 8
8. Compute the moments of healthy longevity from the transient matrix **Ũ**, the transition matrix 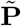, and the reward matrices 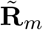, following equations (13) – (15).

## 8 Discussion

The concept of healthy longevity takes it as given that an individual will live for some length of time and will experience various health conditions during that time. The distinction between incidence-based and prevalence-based analyses reflects the information incorporated. Prevalence-based analyses (the Sullivan method and its relatives) incorporate age, but are necessarily blind to the actual transitions among health stages, relying instead on age-specific prevalences. Incidence-based analyses explicitly include transitions among health stages. The great value of prevalence models is that they can be constructed from cross-sectional data, making them an essential tool for exploring healthy longevity when longitudinal data are not available. The analyses are almost always restricted to expectations (health expectancy). Caswell and Zarulli (2018) presented a replacement of the Sullivan method that provides variances and higher moments using MCWR methods, treating prevalence as an age-specific reward.

It is possible, and an interesting exercise, to write prevalence-based models as a degenerate special case of incidence-based analyses. Doing so might bring some extra insight even with limited data; see Appendix D.

Health has many dimensions (physical, mental, emotional, spiritual). It may be defined in terms of specific diseases (e.g., colorectal cancer) or in terms of more general conditions (e.g., disability). It may include combinations of states, some of which are usually described as health while others may be functions of the individual’s actions; e.g., the analysis combining illness and employment by Parker et al. (2020). Longevity can be defined over time scales covering large fractions of the lifetime or for short term conditions (e.g., the model of septic infection by Rangel-Frausto et al. 1998 which describes transitions on a scale of days). For the curious, it is interesting to apply the model to the special cases with only a single health stage (*τ* = 1) or a single age class (*ω* = 1); see Appendix E

We have emphasized the importance of variance (and higher moments) of healthy longevity. In previous studies, analysis of the variance in longevity has been frustrated by the need to analyse sets of states rather than single states. Classical Markov chain theory provides the moments of occupancy for single states, but not for sets of states. The expected value of a sum is the sum of the expected values, but the variance of the sum is not the sum of the variances (except in the special case where the items are independent). The MCWR method we introduce here solves this difficulty, not only for occupancy but for transitions and for values associated with them.^11^

The computational speed of the MCWR calculations will make it easier to use resampling methods (bootstrap, parametric bootstrap, Monte Carlo uncertainty calculations) to estimate the sampling errors of estimates of healthy longevity. It is important to distinguish the variances calculated here, which measure variability among individual outcomes, from variances in estimates of parameters due to sampling error. The former describe a real-world property of the individual or the cohort; the latter describe a property of our knowledge of an estimated quality (Wolf and Laditka, 1997; Salomon et al., 2001).

### 8.1 Notes on the model

The approach presented here solves several important problems in multistate health models.

#### A solution to the inhomogeneity problem

When incidence-based models are written as Markov chains on the set of health states alone, age-dependence renders the Markov chain inhomogeneous, with transition rates that change as a function of age (e.g., Lievre et al., 2003; Cai et al., 2010; van den Hout et al., 2019). Such inhomogeneous systems have no analytical solution. Classifying individuals jointly by age and health stage, as we do here, gives a homogeneous Markov chain on the joint age× state space, as written in (6) and (11). All the usual Markov chain analyses are then available, subject only to the need to keep track of the arrangement of age classes and health stages. The vec-permutation formulation of the model makes this task easy.

#### No limitation to one-way transition patterns

Many incidence-based models in the literature, including the CRC model, contain only one-way, irreversible transitions. This may be due to the irreversibility of some health conditions (e.g., dementia), or it may reflect the attraction of the ability to write the entries of the transition matrix **P** from the intensity matrix **Q** when the transitions are irreversible (e.g., Touraine et al., 2016; Van der Gaag et al., 2015). The availability of reliable routines for computing the matrix exponential makes the latter reason less compelling. Nothing in our analysis is restricted to one-way transitions. If recovery was possible in the CRC model, it would appear as one or more arrows in Figure 1, and as entries in the **Ũ**. The analysis would be the same, and so the model is applicable to conditions from which recovery is possible. Indeed, a possible application of the approach would be to explore the consequences of treatments that would create recovery where none is currently possible.

#### No need for simulations

The transition matrix 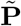 contains all the information needed to carry out individual-based simulations. Following a large number of individuals through their transitions among the transient age-stage combinations until their eventual death produces a sample of the distribution of lives implied by 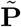. This empirical distribution provides approximate values of means, variances, and other statistics of healthy longevity (e.g., Cai et al., 2010; Wolf and Laditka, 1997).

There are reasons to prefer exact analytical solutions to approximate simulations, especially for exploring alternative scenarios or resampling parameter space, when many calculations are required. Calculating the moments and statistics of longevity in each of the seven states of the CRC model at each of the 50 ages requires less than 0.5 seconds on an ordinary laptop computer. Simulation methods should perhaps be reserved for situations where strong inter-individual interactions make agent-based models essential (e.g., Kluge and Vogt, 2017) or where the entire distribution of healthy longevity is required. Recent results by Tuljapurkar et al. (2020) may, if extended to age×stage-classified models, provide a way to analytically calculate the entire distribution. See (Deconinck et al., 2017) for a comparison of individual simulations and Markov chain models in a study of malnutrition.

#### Independence of estimation methods

The estimation of multistate age×stage-classified models from incidence data is challenging, and a number of methods, tailored to particular kinds of data, are available (e.g., Brouard, 2019; Cai et al., 2010; Lievre et al., 2003; van den Hout et al., 2019; Willekens and Putter, 2014; Willekens, 2014). Some of these methods make strong parametric assumptions (e.g., transition probabilities multiple logistic functions of age) and others are more non-parametric; some provide estimates of continuous-time rates and others estimates of discrete-time probabilities. From the point of view of the MCWR analysis, it makes no difference how the model is estimated. The transition matrix (or the intensity matrix, which is transformed to a transition matrix as in Section 2.1) is all that is needed. The estimation of the matrix is free to proceed in whatever way is most appropriate to the data.

#### A framework for reporting results

There is currently no standard format for reporting multistate models. Every paper seems to do so differently. In our experience most of the literature fails to specify the model clearly enough to permit an evaluation or re-analysis of the results. The theory we have presented here provides a convenient framework in which these models could be presented. Specifying the health stages, the range of age classes, and the transition matrices for each age class would permit any reader to duplicate the analyses. More important, it would permit a reader to make different choices, from among the astronomical number available, of definitions of healthy longevity.

### 8.2 Extensions

The analysis presented here invites extensions. We discuss a few here.

#### Mixed cohorts

The statistics of healthy longevity presented in Figures 3–8 are specific to an individual (or equivalently, a cohort) of a particular initial age and initial health stage. It is sometimes of interest to consider heterogeneous cohorts composed of a mixture of individuals in different health stages. For example, one might ask about the healthy longevity of a cohort of 50-year old individuals distributed among the CRC health stages according to the measured prevalence of those stages at age 50. Let the composition of that mixture be specified by a mixing distribution ***π***, where ***π*** is a probability vector (non-negative entries, summing to 1). The *m*th moments of healthy longevity for this mixed cohort are given by

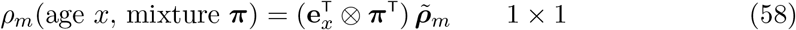

(Caswell et al., 2018; van Daalen and Caswell, 2020). Cohorts comprised of a mixture of ages, or both ages and stages, can be analysed similarly.

#### Variance components

The variance in healthy longevity in a mixed cohort has two components. One is the variance within each health stage, due to individual stochasticity. The other is the variance between health stages, due to heterogeneity in the rates experienced by those individuals. The within- and between-group variance components quantify the extent to which the variability in outcome is due to within-cohort. See (Caswell, 2014; Hartemink et al., 2017) for effects of frailty, (Seaman et al., 2019) for effects of socioeconomic deprivation, and (Hartemink and Caswell, 2018) for latent heterogeneity in laboratory animals. The methodology for multistate models is described in detail in van Daalen and Caswell (2020).

#### Sensitivity analysis

Models are often built not to answer the question “what’s the result?” but the question “how does the result depend on the variables from which it is calculated?” That is the fundamental question of sensitivity analysis (Caswell, 2019). Because the model here is formulated in terms of matrix operations, it is open to sensitivity analysis using matrix calculus, which provides analytical solutions for the derivatives of any model output (scalar-, vector-, or matrix-valued) to any parameters (scalar-, vector-, or matrix-valued). For the theory and many demographic applications see Caswell (2019); for the specific case of multistate age-stage models see Caswell (2012) and Caswell et al. (2018); and for the sensitivity analysis of Markov chains with rewards see van Daalen and Caswell (2017).

#### Other kinds of rewards

The alert reader will have noticed that there is nothing special about the use of “health” to define the stages or the rewards in this model. Indeed, multistate models are widely used to examine a variety of kinds of stages. To the extent that occupancy or transitions or values assigned to them are of interest, the methods we present are directly applicable. See Appendix F for a discussion of applications to lifetime reproductive output, where the rewards are defined in terms of fertility.

If life is a Markov chain, then health is a reward. The combination of this paper and that of Caswell and Zarulli (2018) provide a unified treatment of healthy longevity for incidence-based and prevalence-based models for any kind of health condition.

## Data Availability

All data used in this paper are from publicly available databases.

## 9 Acknowledgments

We gratefully acknowledge financial support from the European Research Council under the European Union’s Seventh Framework Programme (FP7/2007-2013), ERC Advanced Grant 322989 and under the European Union’s Horizon 2020 research and innovation program, ERC Advanced Grant 788195. SvD received support from NWO-ALW Project ALWOP.2015.100. The paper was completed during a Distinguished Lorentz Fellowship from the Netherlands Institute for Advanced Study in the Humanities and Social Sciences (NIAS) awarded to HC. The ideas were developed while teaching courses in health demography at the Max Planck Institute for Demographic Research; we thank the MPIDR for hospitality and the course participants for their interest. Conversations with Mikko Myrskyla, Alyson van Raalte, and Djoeke Schoonenberg, have been helpful. Until the pandemic arrived, Motion Coffee in Amsterdam provided a relaxing environment, for which we are grateful.

## Appendices

There are times when a study leads to interesting or entertaining insights that are not a part of the main story but are too good to lose. In this appendix we present some of these.

### A Matrices unify and simplify demography

Four decades ago, in an issue of the journal Environment and Planning A devoted to multi-state models, Nathan Keyfitz 1980 wrote the following:

Matrices unify and simplify demography. Just as one-dimensional man was declared to be obsolete — by Herbert Marcuse in a book now itself obsolete but of which this one phrase remains — so one-dimensional demography is now transcended by Andrei Rogers and his coworkers.… For many of us, including myself, multidimensionality is esoteric …But we can be sure that what is strange and difficult to us will be natural and easy to our children. What is a narrow and almost closed sect within the profession today is going to standard and obvious demographic technique in the next decade. A table of working life made the way Dublin et al (1949) made theirs, or a table of marriage similarly calculated, will be regarded as quaint; it will compare with the matrix method discussed in this issue as a cumbersome and inflexible desk calculator of the 1940s would compare with a hand-programmable calculator of the late 1970s.

To our children the matrix methods will seem inevitable because they enable demographers to fall back on a hundred years of development in mathematics — due to Sylvester, Frobenius, Kolmogorov and others — rather than to invent the mathematics independently as they go along.

Keyfitz may have been overly optimistic, but his points are nonetheless valid.

### B Calculating the vec-permutation matrix

The vec-permutation matrix is, like any permutation matrix, a square matrix with a single 1 in each row and column and zeros elsewhere. It rearranges, but does not change the value of, the entries in a vector on which it operates. Let **X** be an *m* × *n* matrix,

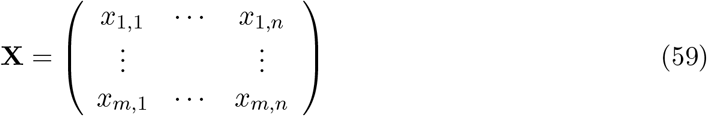

The vec-permutation matrix **K**_*m,n*_ relates the vec of **X** and the vec of **X**^T^:

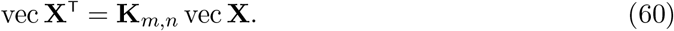

The subscripts on **K** can be omitted if they are clear from the context. The matrix is given by

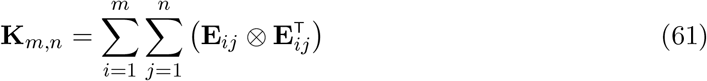

where **E**_*ij*_ is a *m* × *n* matrix with a 1 in the (*i, j*) entry and zeros elsewhere (Henderson and Searle, 1981).

### C Some models for the value of fractional occupancy

In Section 5.3 we presented a model for the reward matrices that specify the value of the partial occupancy of a set of age-stage combinations. We assumed that there was a value for occupancy of an entire interval, with the *m*th moments given by the vector **v**_*m*_, for *m* = 1, …, and that occupancy for a fixed fraction *a* of an interval yields a fixed fraction *a* of the value associated with a full interval (we used *a* = 1*/*2).

Here, we will derive that result and two other possibly interesting models for assigning values to partial occupancy. Define the random variables

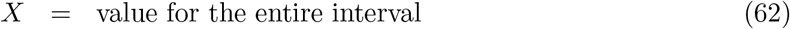

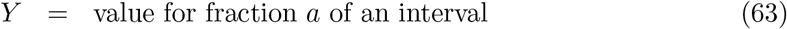

with moment generating functions *M*_*X*_(*s*) and *M*_*Y*_ (*s*). The moment generating function is *M*_*X*_(*s*) = *E* (*e*^*sX*^) and the *m*th moment of *X* is given by

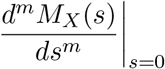

#### Fixed fraction

Suppose that *Y* = *aX*; that is, you receive a fixed fraction *a* of the random reward for occupancy of a full interval. This might apply, for example, for value in terms of annual income.

The moment generating function of *Y* = *aX* satisfies

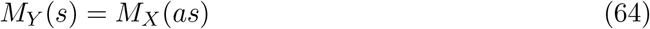

and therefore

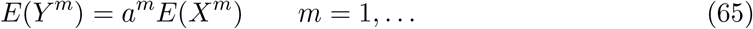

This is implemented in equations (46)–(47).

#### 50-50 chance

Suppose that the value arrives as a packet, or an item, or an event sometime during the interval, and the value of a fraction *a* of an interval is

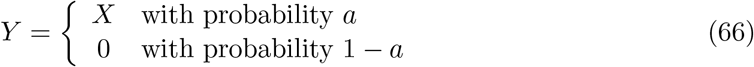

That is, you have a chance *a* or receiving the reward, and a chance 1 − *a* of receiving nothing; examples might include a birth, or a new job.

It is well known that if *Y* is the sum of a random number *N* of independent and identically distributed variables *X*_*i*_,

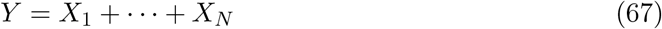

then

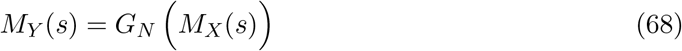

where *G*_*N*_ (*s*) = *E*(*s*^*N*^) is the probability generating function of *N*. In the present case, *N* is a Bernoulli (0-1) random variable with parameter *a*, so

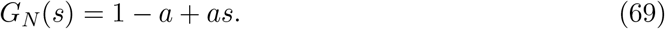

Therefore,

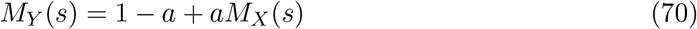

and differentiating gives

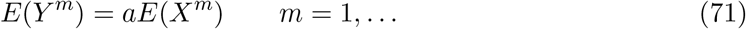

Putting this result into our results for the reward matrices, with *a* = 1*/*2, gives

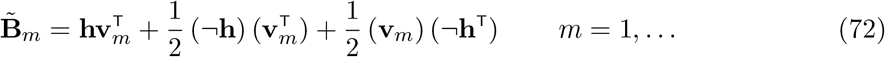

Compare this with (47).

#### Dipping into the stream

Suppose that the value of an interval of occupancy comes from a random stream, or flow, of items arriving as a Poisson process with rate parameter *λ*. The accumulated value *X* of this flow over a unit interval has a Poisson distribution *X* ∼ Poisson(*λ*) with *E*(*X*) = *λ*. Now suppose that the flow operates for only a fraction *a* of an interval. The accumulated value at this point is *Y* ∼ Poisson(*aλ*).

Repeatedly differentiating the moment generating function, or looking up the moments of the Poisson distribution, yields

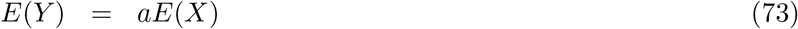

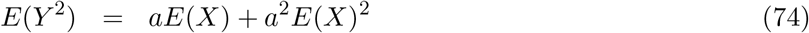

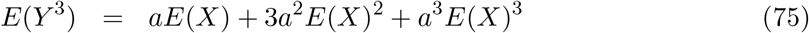

and so on. There is no closed form expression for these moments.

In terms of the reward matrices 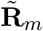, this leads to

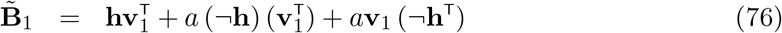

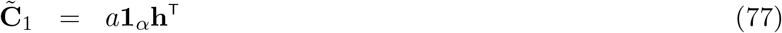

and then

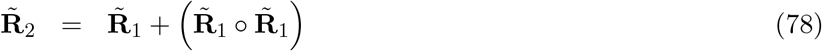

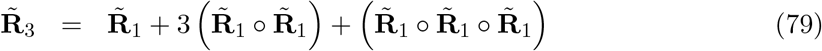

### D Prevalence is a special case of incidence

The calculation of health expectancy from prevalence data by the Sullivan method relies on two strong assumptions. The first is that the chance of being healthy at age *x*, which is given by the prevalence of being healthy at age *x*, is independent of the health states at *x* − 1 or *x* + 1. The second is that the probability of survival from age *x* to age *x* + 1 is independent of the health state at *x*. These assumptions are strong but unavoidable in the absence of longitudinal individual data. It’s not the fault of the method.

However, independence is a special case of dependence, so it is interesting to deploy our theory in a prevalence-based analysis. Consider the two-state model shown in Figure 10. Let *v*_1_(*x*) and *v*_2_(*x*) be the prevalences of the healthy and not-healthy states at age *x*, where *v*_1_ + *v*_2_ = 1, and let *p*(*x*) be the survival probability from age *x* to *x* + 1. Then the transition matrix **U**_*x*_ is

**Figure 10:**
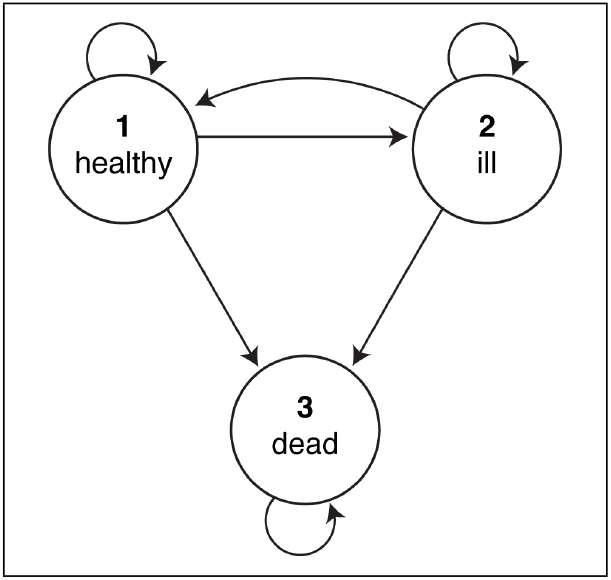
Transition graph for a two-stage health model (healthy and not-healthy) with death as an absorbing state.

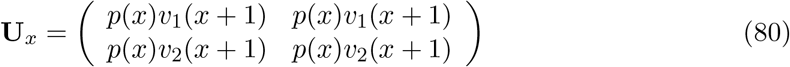

The equality of the columns of **U**_*x*_ means that the health state at *x* + 1 is independent of the health state at *x*. The survivors from age *x* (with probability *p*(*x*)) are allocated to healthy and not healthy states according to the prevalences at *x* + 1. The transition matrix for the multistate Markov chain is 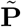 as given by equation (11). The MCWR approach then applies directly to this model, with the reward matrices calculated from the array **H** exactly as in Section 5.1. This makes prevalence-based calculations more flexible in the choice of ages over which to compute healthy longevity. And, of course, it provides not only the mean, but all the moments of healthy longevity.

It is apparent from this formulation how restrictive the hypotheses of prevalence-based analysis are. However, it also suggests some modifications to relax the assumptions. It is simple to extend the number of health stages (e.g., healthy, slightly ill, severely ill, etc.). It is also possible to incorporate differential survival as a function of health state, if such information is available. For example, suppose there are three health stages, and let *v*_*i*_(*x*) be the prevalence of health stage *i* at age *x*, where Σ _*i*_ *v*_*i*_(*x*) = 1. Let *p*_*i*_(*x*) be the survival probability of an individual in health stage *i* at age *x*. Then the matrix **U**_*x*_ becomes,

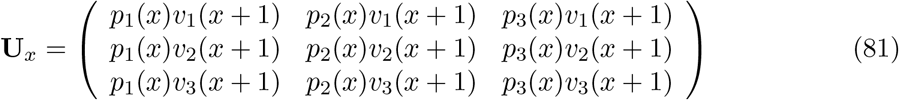

Now the health state at *x* + 1 is independent of the health state at *x*, but the survival probability is not. The multistate transition matrix 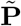 is constructed as usual, and the array **H** can now be used to compute healthy longevity in any desired combination of age classes and health stages.

We invite researchers confronted with prevalence data to use this analysis as a complement to the MCWR analysis of Caswell and Zarulli (2018) and as an alternative to the much more limited Sullivan method.

### E One is a special case of many

We note that one is simply a special case of many; so we can apply the analysis to the very special case of only a single stage or only a single age class. This is as simple as setting *τ* = 1, or *ω* = 1, or both.

#### A single transient stage

*τ* = 1, *ω* > 1. Suppose there is only a single way to be alive (or to be present, if exit from the transient stage is not death, but emigration, or graduation, or divorce, etc.). This is the model underlying classical survival analysis, dependent only on age.

Suppose for convenience that *ω* = 3. Then

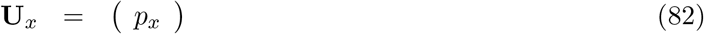

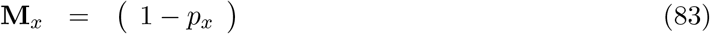

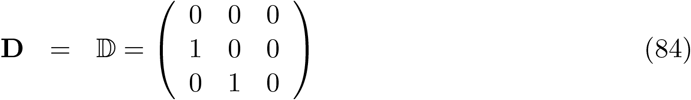

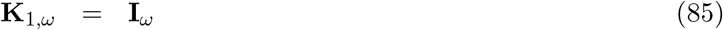

where *p*_*x*_ is the survival probability of age class *x*. The result is

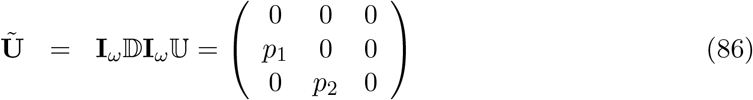

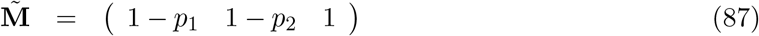

The matrix **Ũ** is, as expected, an age-classified Leslie survival matrix.

The health occupancy matrix **H** is 1 × *ω*. If longevity is measured by lifetime occupancy of the living stage.^12^ Then

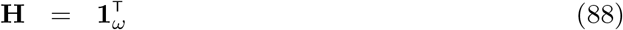

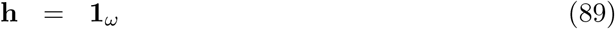

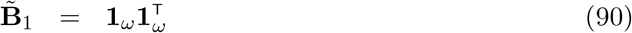

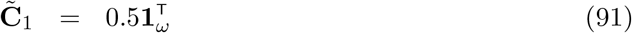

In this situation, the fundamental matrix is known to be

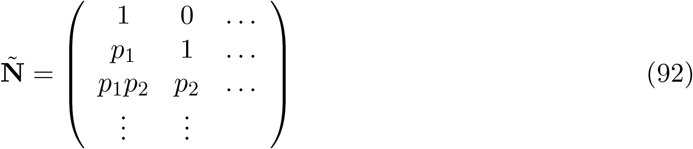

(Keyfitz and Caswell, 2005, Chap. 10). The result of evaluating 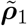 using (13) for this special case is equivalent to the column sums of **Ñ**, minus 1*/*2 to account for the fractional occupancy of the alive stage at the time of death.

#### A single age class

*τ* > 1, *ω* = 1. This is equivalent to asserting that age has no effect on individual fates, which are completely determined by the stage. It is applicable not only to health models, but to stage-classified demographic models common in animal and plant demography (e.g., Caswell, 2001; Salguero-Gómez et al., 2015).

This model includes a single transient matrix **U**, and

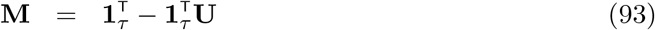

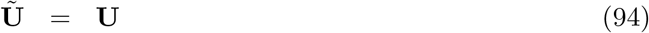

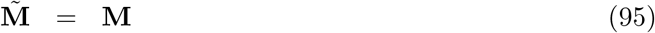

The health occupancy matrix **H** is a 0-1 vector of dimension *τ* × 1, with ones indicating the stages whose occupancy is to be calculated. The analysis gives all the moments of the occupancy time for the chosen set of states.

#### No ages, no stages

*τ* = *ω* = 1. In this case age has no effect on transitions and all individuals are identical and not differentiated by stage. This is the model for the decay of radioisotopes. The atoms are all identical (so there are no stages) and nothing is affected by how long the atom has been there. The matrices take particularly simple forms

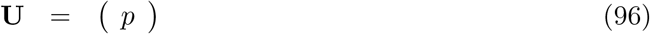

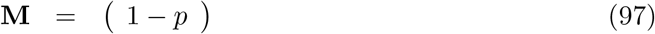

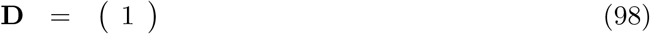

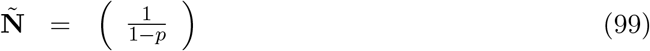

and **Ũ** = **U**, 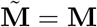, and

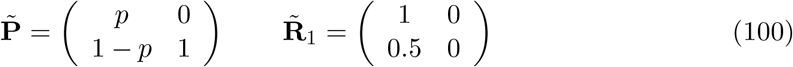

Substituting these into the expression (13) yields

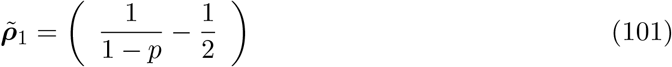

The differs by 1/2 from the estimate of life expectancy obtained by the column sum of **Ñ**. That sum is the mean of the geometric distribution of the number of trials before absorption; the difference reflects a decision about how to turn trials into time units.

### F Reproduction is special case of value

Lifetime reproductive output (abbreviated LRO) is the total number of offspring produced over the life of an individual. LRO is a component of fitness, and variance in LRO plays an important role in evolutionary demography (Crow, 1958; Brown et al., 2009; van Daalen and Caswell, 2020).

The mean of LRO is the net reproductive rate *R*_0_. But LRO, like longevity, is stochastic, because of randomness in the path through the life cycle (how long, and through what sequence of stages, does the individual live?) and in the amoung of reproduction at each age or each age-stage combination. The moments of LRO, including both sources of randomness, can be calculated using Markov chains with rewards (Caswell, 2011; van Daalen and Caswell, 2015; van Daalen and Caswell, 2017).

The method presented here for assigning values to the occupancy of a state or transitions among states, as in Sections 5.3 and 5.4, is directly applicable to LRO. Setting **V**_*m*_(*i, j*) in (44) to the *m*th moment of reproductive output at stage *i* and age *j*. The value in **V**_*m*_(*i, j*) in equation (44) would be the *m*th moment of the amount of reproduction associated with a unit of occupancy of stage *i* at age *j*. For humans (and monovular animals like whales), if multiple births are ignored, reproduction is a Bernoulli (0 − 1) random variable. For other species, other distributions are appropriate. The resulting calculation of lifetime accumulated value gives the LRO for the set of age-stage combinations in the set 𝒮. This provides a completely flexible definition of LRO, including possibilities like early LRO and late LRO, defined in the same way as early and late cancer conditions in the CRC example.

The death of an individual results not only in the loss of a certain amount of longevity, but also of a certain amount of future reproduction. Replacing the vectors 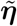 in equation (56) with the appropriate vectors of the moments of LRO would give the moments of lifetime reproduction lost due to a specified cause of death.

Incorporating LRO into multistate models including health stages would provide a way to explore health effects on fertility. Diabetes, for example, has well known negative effects on fertility (e.g., Wiebe et al., 2014; Lin et al., 2018). Such results could, in principle, be used to develop appropriate reward functions to be combined with an incidence-based diabetes model like that of Kuo et al. (1999) to evaluate the health consequences for fertility.

Less directly health-related, but of great interest for studies of fertility, would be multi-state MCWR models including marital status (e.g., Keyfitz, 1988) or family and partnership structure (Mills, 2000) as stages. Caswell (2020) presented an age×parity-classified model for the analysis of kinship; it could be analysed for LRO using the methods here. A model based on age and parity is interesting because reproduction is associated not with occupancy, but with the transition, from one parity class to the next.^13^ A model in which stages are defined by maternal age (i.e., the age of the mother at the birth of the individual) has been developed by Hernández et al. (2020) and applied to senescence in the rotifer *Brachionus manjavacas*, a model organism used in the study of ageing. van Daalen et al. (2020) have extended the model to examine the statistics of LRO, an approach that could be extended in general the late life consequences of early life conditions.

## Footnotes

1 It is not hard to find statements to this effect. “Such a deterministic forecast, however, does not give an accurate view of forecast uncertainty. The future is inherently uncertain, and hence probabilistic methods have to be used.” (Christiansen and Keilman, 2013). Or, as a Governor of the Bank of England put it, “An informed discussion of public policy issues, however, requires an analysis of the risks and uncertainties involved. Whether in policies for health or transport, matters monetary or meteorological, in times of war and peace, decisions should reflect a balance of risks. Yet policy debates continue to be permeated by the ‘illusion of certainty.’ “ (King, 2005).

2 It is best not to be distracted by concerns with non-Markovian dependence. Such phenomena usually mean that the state space of the model has yet to be properly defined.

3 The Kronecker product of a *m × n* matrix **A** and a *p × q* matrix **B** is the *mp × nq* matrix 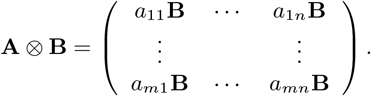

4 An alternative commonly used approximation requires, in addition to the assumption of constancy, the assumption that the elements of **Q** are sufficiently small that a first-order approximation is suitable (Rogers and Ledent, 1976). For transitions over an interval of length Δ*t*, the approximation is 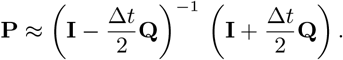 See Schoen (1988) for a comparison of approaches to deriving discrete transition matrices from continuous rate matrices.

5 Sources of mortality can, if desired, also be incorporated into the **D***j*. See Caswell et al. (2018) for details.

6 A alternative formulation was given by Roth and Caswell (2018). The approach we use here is somewhat simpler and more flexible.

7 The notation here differs from that in van Daalen and Caswell (2017). In that paper, the tilde on 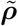 denoted a vector from which the absorbing states had been removed. Here, however, the tilde denotes vectors and matrices with the block structure of stages within age classes, as given by (6).

8 It is understood that interest might focus on longevity subject to some illness or condition that is not pleasant or desirable.

9 In this model, it is impossible to leave the clinical CRC stages, other than by death. Counting the transitions might be of limited interest, but it is easy to imagine cases where transitions would be more flexible.

10 This is not an ideal example of a YLL calculation because, in the model of Wu et al. (2006), the transition rate from clinical CRC to death due to CRC does not change with age, whereas the rate of mortality due to other causes does. But it is useful as an example.

11 Roth and Caswell (2018) showed how to extract occupancy of sets of states by a transformation of the transition matrix **P**, but the result has yet to be extended to multistate models, to transition, or to values associated with occupancy and transitions.

12 Note that it is possible to consider occupancy of the living stage at some subset of age classes of interest.

13 Similar considerations apply to colonially nesting seabirds, where reproduction is associated with a return to the breeding colony (e.g., Jenouvrier et al., 2005).

